# Beyond Metrics to Methods: A Scoping Review of Large Language Models for Detection of Social Drivers of Health in Clinical Notes

**DOI:** 10.1101/2025.07.04.25330866

**Authors:** Ahmed Farrag, Ahmed Soliman, Elham Hatef, Amie Goodin, Masoud Rouhizadeh

## Abstract

**Objective:** This scoping review aimed to map current applications of Large language models (LLMs) for extracting Social drivers of health (SDoH), benchmarks model performance across domains to define the state of the field, and evaluates methodological approaches to identify research gaps and guide clinical deployment.

**Materials and Methods:** We searched PubMed, Web of Science, Embase, Scopus, and IEEE Xplore for studies applying LLMs in the detection of SDoH. We applied a novel methodological framework integrating: (1) a hierarchical classification system for SDoH domains and LLM architectures; (2) a systematic approach for synthesizing performance metrics; and (3) a custom seven-domain instrument to assess the methodological rigor.

**Results:** Forty-two studies met inclusion criteria. Behavioral Factors had the highest median F1-score (0.87), while Health Care Access and Quality showed the lowest and most variability (median F1 = 0.59). Research was concentrated in the United States (85.7%) and private institutional datasets (69%), often focused on critical care populations (45.2%). Methodological assessment revealed that only 29% of studies provided annotation guidelines, 24% assessed fairness across demographic groups, and 21% validated models externally.

**Discussion and Conclusion:** The progress of using LLMs for SDoH extraction is limited by performance variability, weak methodological rigor in the conducted studies, and minimal attention given to fairness and generalizability. Methodological gaps include a lack of provided annotation guidelines, assessment of fairness, and external model validation. LLMs show strong potential for extracting SDoH from clinical text. However, to move forward, addressing the current limitations demands more standardized, transparent, and robust research.

## 1. BACKGROUND AND SIGNIFICANCE

Social drivers of health (SDoH)—defined by the World Health Organization (WHO) as the non-medical factors influencing health outcomes—determine an estimated 70-80% of modifiable health contributions across populations.^1–3^ A robust body of research, including longitudinal cohort studies and health disparities analyses, consistently demonstrates that socioeconomic status, environmental exposures, and psychosocial factors exert a greater impact on population health than clinical care alone.^4–9^ Despite these well-established associations, healthcare systems continue to face major challenges to systematically capture, document, or integrate these critical determinants into routine clinical practice, creating a fundamental disconnect between epidemiological evidence and care delivery.^10^

Although electronic health records (EHRs) have transformed healthcare documentation, they remain limited in their capacity to capture SDoH.^11^ Analyses of national Medicare claims indicate that structured SDoH codes (ICD-10-CM Z-codes Z55–Z65) are used in fewer than 1.5% of patient encounters, largely due to clinician time constraints, a lack of standardized screening protocols, and limited integration into care pathways.^12,13^ In contrast, natural language processing (NLP) studies estimate that 83–91% of SDoH documentation resides within unstructured clinical narratives— representing both a rich source of untapped information and a significant informatics challenge.^14^

Three fundamental barriers impede effective SDoH extraction from clinical text: (1) documentation heterogeneity, characterized by low prevalence and high linguistic variability of SDoH mentions across providers and settings; (2) architectural constraints within EHR systems that limit SDoH data capture and interoperability; and (3) analytical limitations of traditional NLP techniques, which lack the contextual understanding required to identify implicit social determinants.^15–18^ Early rule-based and conventional machine learning approaches, though interpretable and relatively simple to deploy, failed to capture subtle, context-dependent language pervasive in clinical documentation.^19,20^ Identifying housing instability, for instance, requires systems to recognize that phrases like “patient sleeps on friend’s couch” or “lives on streets” signal homelessness—inferences that require contextual understanding beyond simple keyword matching.^21^

Large language models (LLMs) represent a paradigmatic shift in clinical NLP capabilities for SDoH extraction. These transformer-based neural architectures leverage massive pretraining corpora to develop nuanced linguistic understanding, enabling superior performance on complex extraction tasks despite limited labeled data.^22–25^ Encoder-based architectures (BERT, ClinicalBERT, BioBERT) excel at classification through bidirectional context modeling, while decoder-based models (GPT variants) demonstrate few-shot learning capabilities that address data scarcity challenges endemic to SDoH research.^22–25^ Recent studies suggest that models such as BERT and GPT-4 outperform earlier methods—and, in some cases, even human reviewers—in detecting implicit or rare SDoH within clinical narratives.^26–28^

Despite these advances, critical methodological and ethical challenges constrain the clinical deployment of LLMs for SDoH extraction. Current implementations are often fragmented with inconsistent performance metrics, varied validation practices, and limited methodological transparency. This methodological heterogeneity prevents meaningful cross-study comparisons and evidence synthesis needed to establish clinical best practices. More concerning, pre-trained models inherit societal biases from their training corpora, potentially perpetuating or amplifying health inequities when applied to marginalized populations—a fundamental ethical challenge given that SDoH applications specifically target vulnerable groups requiring equitable representation and accurate identification.^29–32^

## 2. OBJECTIVE

This scoping review aimed to: (1) systematically map the SDoH domains and LLM architectures investigated to identify research gaps; (2) establish a standardized benchmark of LLM performance across SDoH domains and subdomains to serve as a field-level reference for future evaluations; (3) compare model performance using quantitative metrics; and (4) assess methodological rigor across internal validity, external validity, and reporting transparency. By integrating both performance outcomes and methodological appraisal, this review provides a dual-level benchmark to guide the development of robust and clinically meaningful LLM applications for SDoH extraction.

## 3. MATERIALS AND METHODS

### 3.1. Study Design

We conducted a scoping review following the Preferred Reporting Items for Systematic Reviews and Meta-Analyses Extension for Scoping Reviews (PRISMA-ScR) checklist and the Enhancing Transparency in Reporting the Synthesis of Qualitative Research (ENTREQ) guidelines.^33,34^ Full documentation of these frameworks is provided in the Supplementary Materials; **PRISMA-ScR Checklist** and **Appendix A.**

### 3.2. Databases and Search Strategy

To comprehensively capture literature at the intersection of clinical medicine, informatics, and machine learning, we searched five electronic databases: PubMed/MEDLINE, Web of Science, Embase, Scopus, and IEEE Xplore. Search strategies were collaboratively developed by a medical informaticist and an information specialist experienced in systematic reviews. Queries were tailored for each database using both controlled vocabulary (e.g., Medical Subject Headings [MeSH]) and free-text terms and structured around three core concepts: (1) large language models (LLMs), such as “GPT,” “BERT,” and “transformer architecture”; (2) social determinants or drivers of health (SDoH), including “health equity,” “food insecurity,” and “housing instability”; and (3) clinical free-text sources, such as “electronic health record (EHR) narratives” and “clinical notes.” The initial search was conducted on January 15, 2025, and updated on March 1, 2025. No publication date restrictions were applied. Complete search strategies for all databases are available in **Supplementary Appendix B, Section 1**.

### 3.3. Eligibility Criteria

We included studies that met four criteria: (1) analyzed clinical free-text data—including EHR notes or synthetic clinical narratives simulating real-world documentation—for SDoH as defined by the World Health Organization (WHO), U.S. Department of Health and Human Services (HHS), Centers for Disease Control and Prevention (CDC), or Centers for Medicare & Medicaid Services (CMS);^2,35,36^ (2) applied LLMs—whether pre-trained, fine-tuned, or instruction-tuned models such as GPT, BERT, or Flan-T5—for SDoH information extraction tasks; (3) reported quantitative performance metrics (e.g., F1-score, precision, recall) relevant to clinical natural language processing tasks such as named entity recognition, concept extraction, or phenotyping; and (4) presented original research with sufficient methodological detail, including peer-reviewed publications or high-quality preprints.

We excluded studies that met any of the following criteria: (1) used only traditional machine learning methods (e.g., logistic regression, support vector machines) or rule-based systems without comparison to an LLM; (2) focused solely on structured data (e.g., International Classification of Diseases [ICD] codes) or non-clinical sources (e.g., surveys, social media); (3) consisted of review articles, editorials, opinion pieces, or preprints lacking methodological transparency; (4) did not report SDoH-specific performance metrics or presented only qualitative findings; and (5) reported performance as aggregated macro or micro averages across SDoH categories without disaggregated domain-level metrics.

### 3.4. Study Selection Process

All citations were imported into Mendeley (version 1.19.8) and deduplicated. Before screening, a random subset of 10 studies was independently reviewed by all team members to calibrate decision criteria, achieving strong inter-rater agreement (Cohen’s kappa > 0.8). Two reviewers (A.F. and A.S.) independently screened all titles and abstracts using Covidence (Veritas Health Innovation). Conflicts were resolved through discussion. Articles that passed the initial screening underwent full-text review by the same reviewers, with final inclusion decisions made by consensus. The selection process was documented using the PRISMA 2020 flow diagram.

### 3.5. Data Extraction and Synthesis

To analyze and compare heterogeneous LLM-based approaches to SDoH extraction, we developed a structured methodological framework (**Figure 1**; described in Sections **3.5.1** to **3.5.3**). This framework supported standardized extraction of five core data categories: study characteristics, model architecture and implementation, dataset attributes, SDoH classification, and performance metrics. Two reviewers (A.F. and A.S.) piloted the extraction form on 10 studies and independently extracted data from all included articles. A third reviewer (M.R.) validated a random 20% subset, stratified by publication year, achieving substantial agreement (Cohen’s kappa = 0.92). Discrepancies were resolved through structured adjudication or senior arbitration (M.R.).

**Figure 1.**
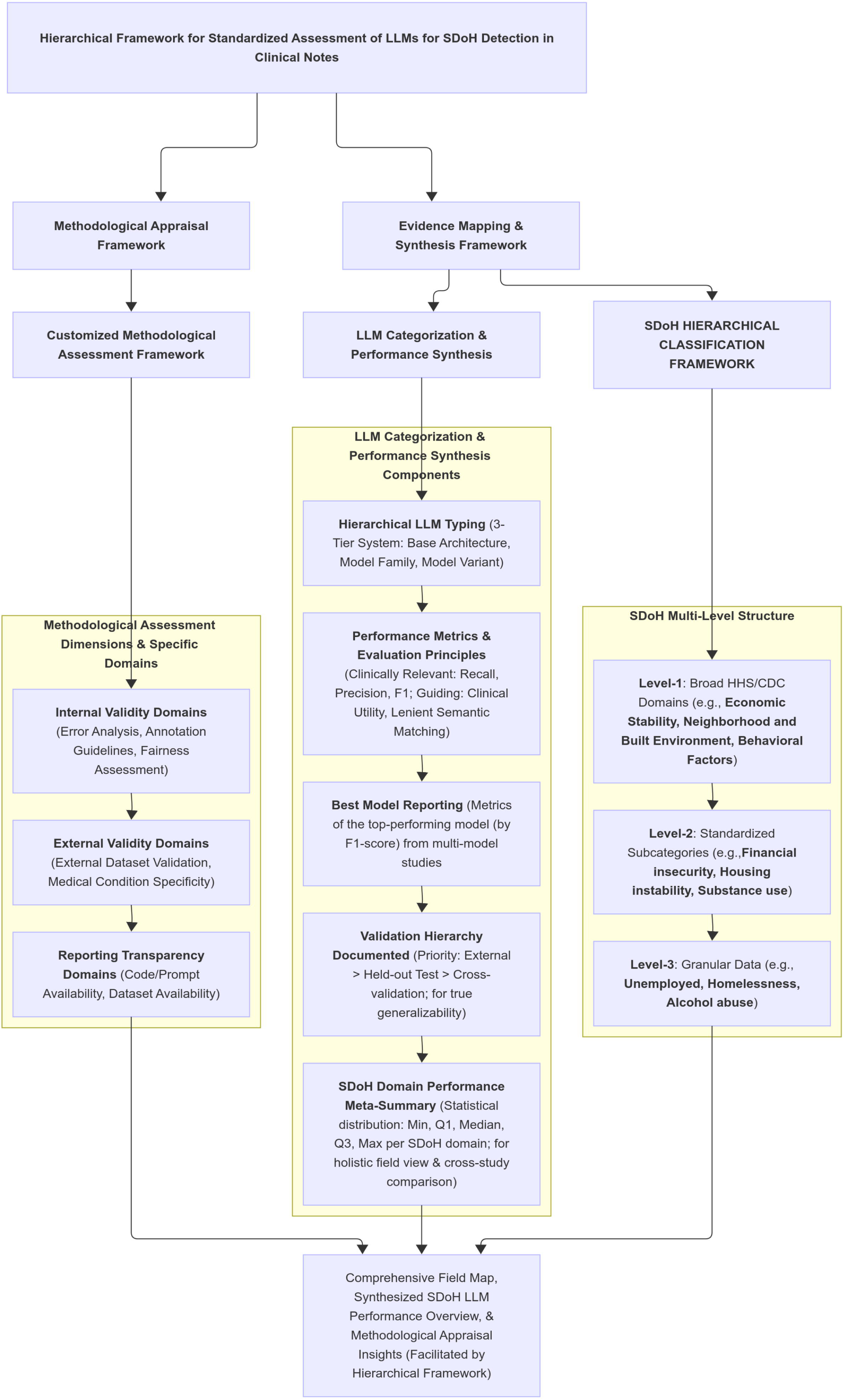
Hierarchical Framework for Standardized Evaluation of LLMs in SDoH Extraction from Clinical Text. This flowchart presents the dual-pronged methodological framework applied in the review. The first component is a *Methodological Appraisal Framework* used to evaluate study quality across three domains: internal validity, external validity, and reporting transparency. The second is an *Evidence Mapping & Synthesis Framework*, which standardizes comparison of studies by hierarchically classifying both Large Language Models (LLMs) and Social Drivers of Health (SDoH), and systematically aggregating reported performance metrics. Together, these components support a comprehensive synthesis of LLM performance and methodological rigor across the field.

#### 3.5.1 SDoH Classification and Performance Framework

We adopted the U.S. HHS Healthy People 2030 framework as the primary taxonomy for SDoH, supplemented by the WHO Commission on Social Determinants of Health and CDC domains.^2,3,35^ These frameworks organize SDoH into five domains: Economic Stability, Education Access and Quality, Health Care Access and Quality, Neighborhood and Built Environment, and Social and Community Context. Additionally, we included Behavioral Factors (e.g., substance use, physical activity, and nutrition) as a sixth independent domain to reflect their clinical importance, modifiability, and documentation prevalence in EHR notes.^2,3,35,37–39^

SDoH performance metrics were aggregated at the level-2 (subcategory) hierarchy.^40^ This allowed us to balance analytic feasibility with granularity and to harmonize reporting across studies with variable annotation detail. When studies reported multiple attributes for the same factor (e.g., “current smoker” vs. “former smoker”), we prioritized extraction of current patient-specific status for consistency and clinical relevance. Broader attributes such as duration, severity, and family history were also recorded and documented in the **Supplementary Materials (Appendix B)** for comprehensive analysis.

#### 3.5.2 LLMs Categorization and Performance Metrics

We classified each model into a three-tier hierarchy: base architecture (e.g., T5), model family (e.g., Flan-T5), and specific variant (e.g., Flan-T5-Large).^41^ For studies that evaluated multiple models, we adopted the more robust approach of selecting a single, overall best-performing model and extracting all its associated metrics. To this end, from each study presenting multiple models, we identified the ‘best-performing model’ as the one achieving the highest reported F1-score, given that the F1-score comprehensively balances recall and precision. All reported metrics (recall, precision, and F1-score) for this single, selected model were then recorded for our analysis. For example, consider a study comparing three models: Model A: Recall = 0.70, Precision = 0.80, F1-score = 0.75; Model B: Recall = 0.85, Precision = 0.75, F1-score = 0.79; Model C: Recall = 0.80, Precision = 0.82, F1-score = 0.81. In this scenario, Model C would be selected as the best-performing model due to its superior F1 score (0.81). Consequently, we would record Model C’s recall (0.80), Model C’s precision (0.82), and Model C’s F1-score (0.81) for our analysis. This method ensured that our evaluation reflected the holistic performance capabilities of a single, optimized model from each study, rather than an amalgam of potentially unrelated peak scores across different models for individual metrics.^42^

To reflect practical clinical utility, we favored lenient over strict span-matching metrics when available, acknowledging the variability of real-world SDoH documentation in clinical notes.^43^ We also applied a validation hierarchy to contextualize generalizability: external validation on independent datasets was considered the gold standard, followed by held-out testing, and cross-validation.^44^ Domain-level summary statistics (e.g., median, interquartile range) were calculated for each SDoH category, with finer-grained metrics reported in the **Supplementary Materials - Appendix B.**

### 3.6. Methodological Assessment

Recognizing the absence of established guidelines for evaluating LLMs in SDoH extraction, we developed a structured assessment framework spanning three key dimensions: internal validity, external validity, and reporting transparency. These dimensions were informed by patterns identified during preliminary review and are designed to reflect both technical rigor and clinical relevance (**Table 1**).^45–52^

**Table 1.**
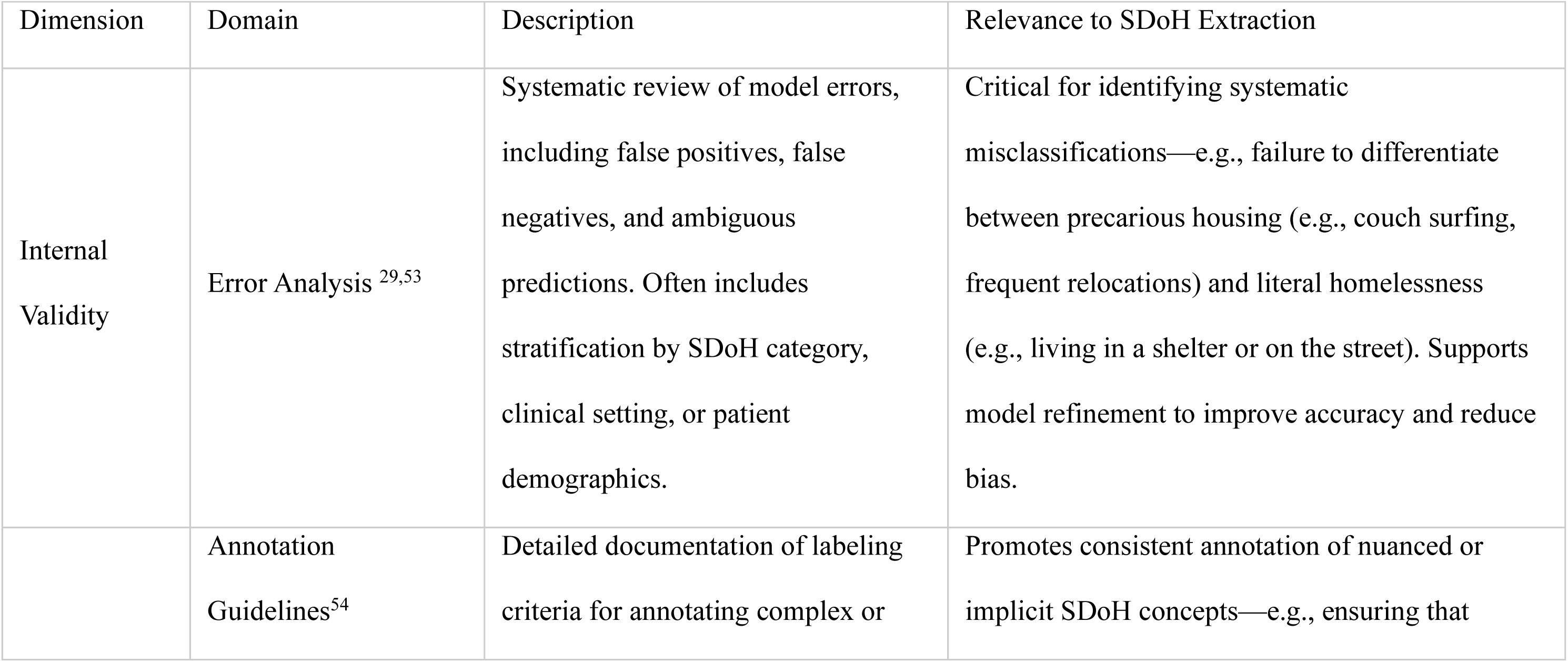
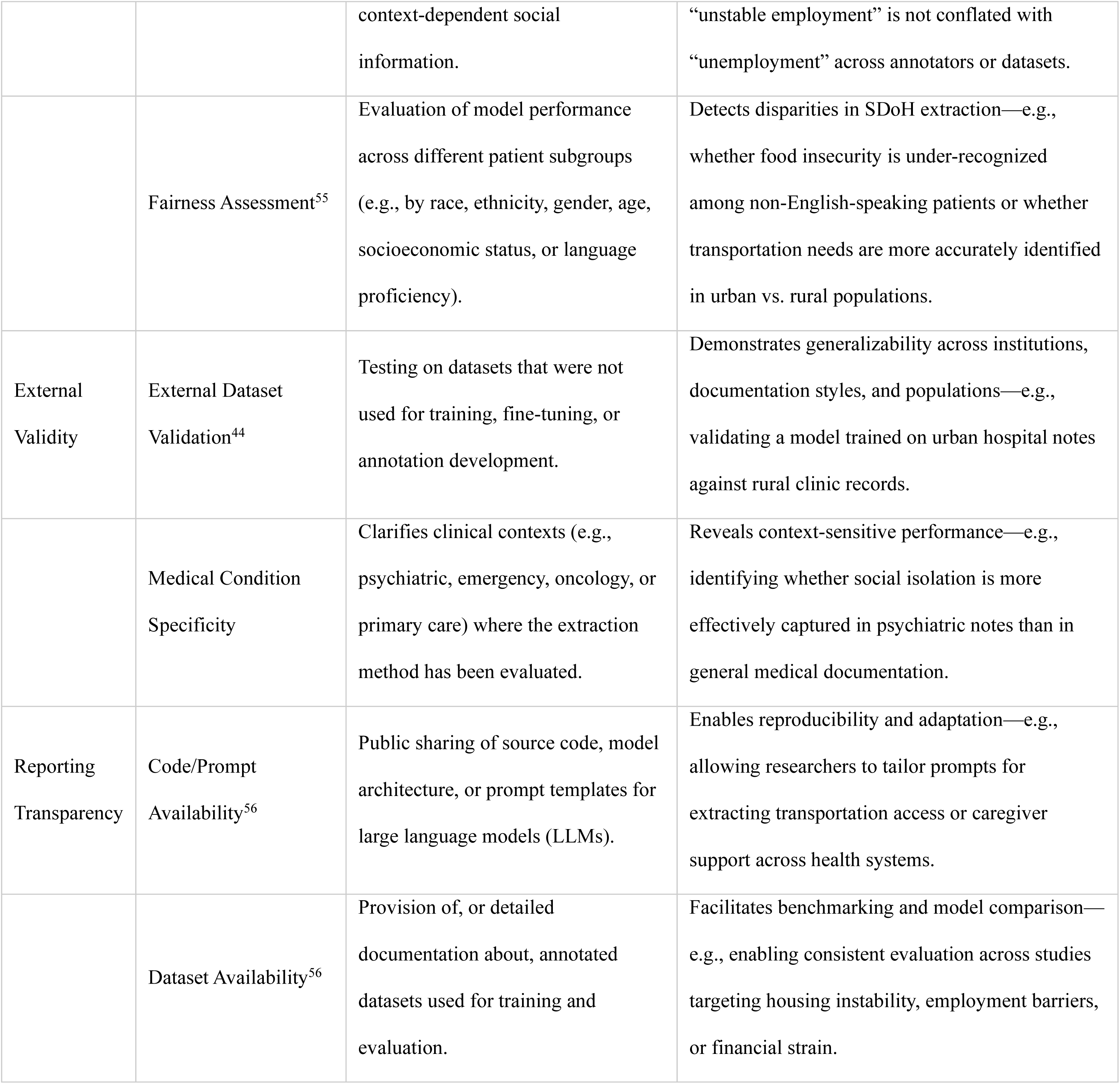
Methodological Domains Assessed in this Review.

Internal validity was assessed through three domains: annotation guidelines, error analysis, and subgroup fairness. We examined whether studies documented clear labeling criteria, conducted error analyses to identify model limitations (e.g., conflation of couch surfing with literal homelessness), and evaluated model performance across demographic subgroups.^29,53^ External validity focused on whether models were tested on independent datasets and the clinical specificity of the documentation analyzed.^44^ Reporting transparency included availability of code, prompts, and datasets, which are essential for reproducibility.^56^ Full rationale for domain selection is provided in Supplementary Appendix C.

Although this framework is not a validated instrument, it offers a pragmatic structure for assessing the current evidence base and identifying methodological gaps. It supports consistent evaluation of LLM-based approaches to SDoH extraction and fosters methodological refinement in this emerging field.

## 4. RESULTS

From 254 records retrieved across Scopus, Embase, PubMed, and IEEE Xplore (through March 1, 2025), 177 titles and abstracts were screened after 95 duplicates were removed. Eighteen additional studies were identified via reference snowballing. Of the 132 full texts assessed, 42 met inclusion criteria ^26,27,64–73,28,74–83,57,84–93,58,94,95,59–63^ (**Supplementary Table**). The main reasons for exclusion were reliance on structured/non-clinical text, use of traditional ML only, or unrelated clinical focus (**Figure 2**).

**Figure 2.**
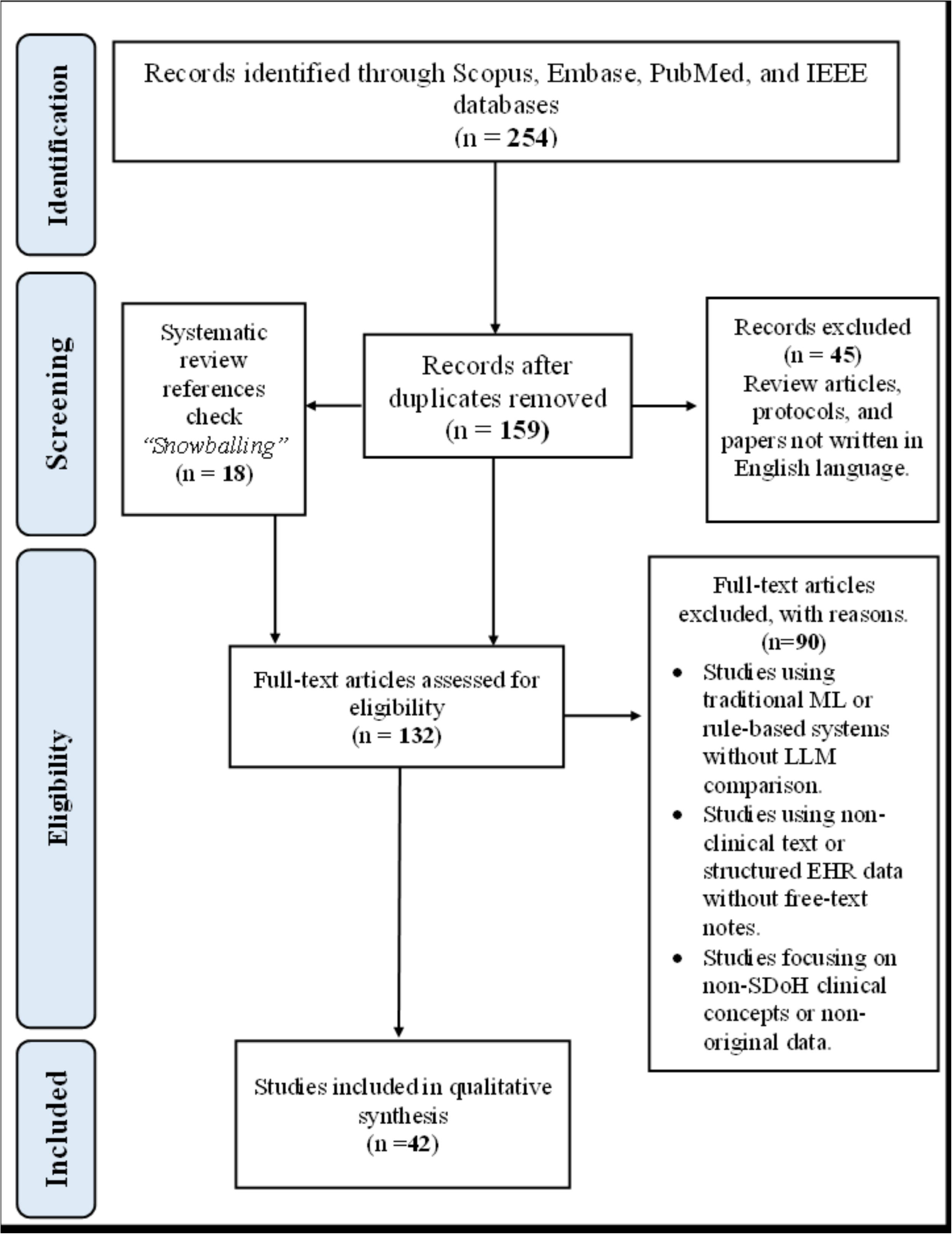
PRISMA 2020 Flow Diagram for Study Selection. This diagram outlines the study identification and selection process. A total of 254 records were retrieved from four databases, with 159 remaining after duplicate removal. An additional 18 records were identified through reference snowballing. Following full-text screening of 132 articles, 42 studies met the inclusion criteria and were retained for qualitative synthesis. *Abbreviations: EHR, electronic health records; ML, machine learning; LLM, large language model; SDoH, Social Drivers of Health*.

### 4.1. Data Sources and Patient Characteristics

We observed considerable data heterogeneity across the included studies (**Appendix B, Section 2**). Most studies (33/42; **79%**) used private institutional datasets, which limits reproducibility. Public datasets appeared in 9 studies (**21%**), with MIMIC-III or MIMIC-IV utilized in 8 out of 9 public cases, reflecting a pronounced bias toward critical care contexts.

Dataset sizes ranged widely, from as few as 100 notes to as many as 50,000, underscoring substantial variation in training data magnitude. This spectrum also illustrates the evolution from BERT-like models, which often require large annotated corpora, to GPT-based or retrieval-augmented models—many of which achieve competitive results with relatively limited examples.

Geographically, the studies were overwhelmingly U.S.-based (**83%**, 35/42), with limited representation from Korea, Spain, France, Austria, and the UK. This U.S.-centrism further constrains the global generalizability of model findings.

Sampling was predominantly convenience-based. Nearly half of studies (**19/42; 45%**) focused on critical care populations, largely driven by the widespread use of MIMIC datasets. Oncology (5/42, 12%), cardiology (4/42, 10%), and psychiatric cohorts (4/42, 10%) were also examined, while pediatric, transplant, and other subpopulations were infrequently represented. As a result, generalizability across disease areas and patient groups remains limited and poorly characterized. A full description of datasets and study populations is available in **Appendix B** (**Section 2**).

### 4.2. Performance of LLM Variants Across SDoH Categories and Subcategories

Across the six major SDoH domains, LLM performance exhibited considerable variability, with marked differences in effectiveness depending on the specific subdomain and metric evaluated (**Figure 3**). Behavioral and social domains yielded the strongest and most consistent results, while health care access and mental health remained persistent challenges.

**Figure 3.**
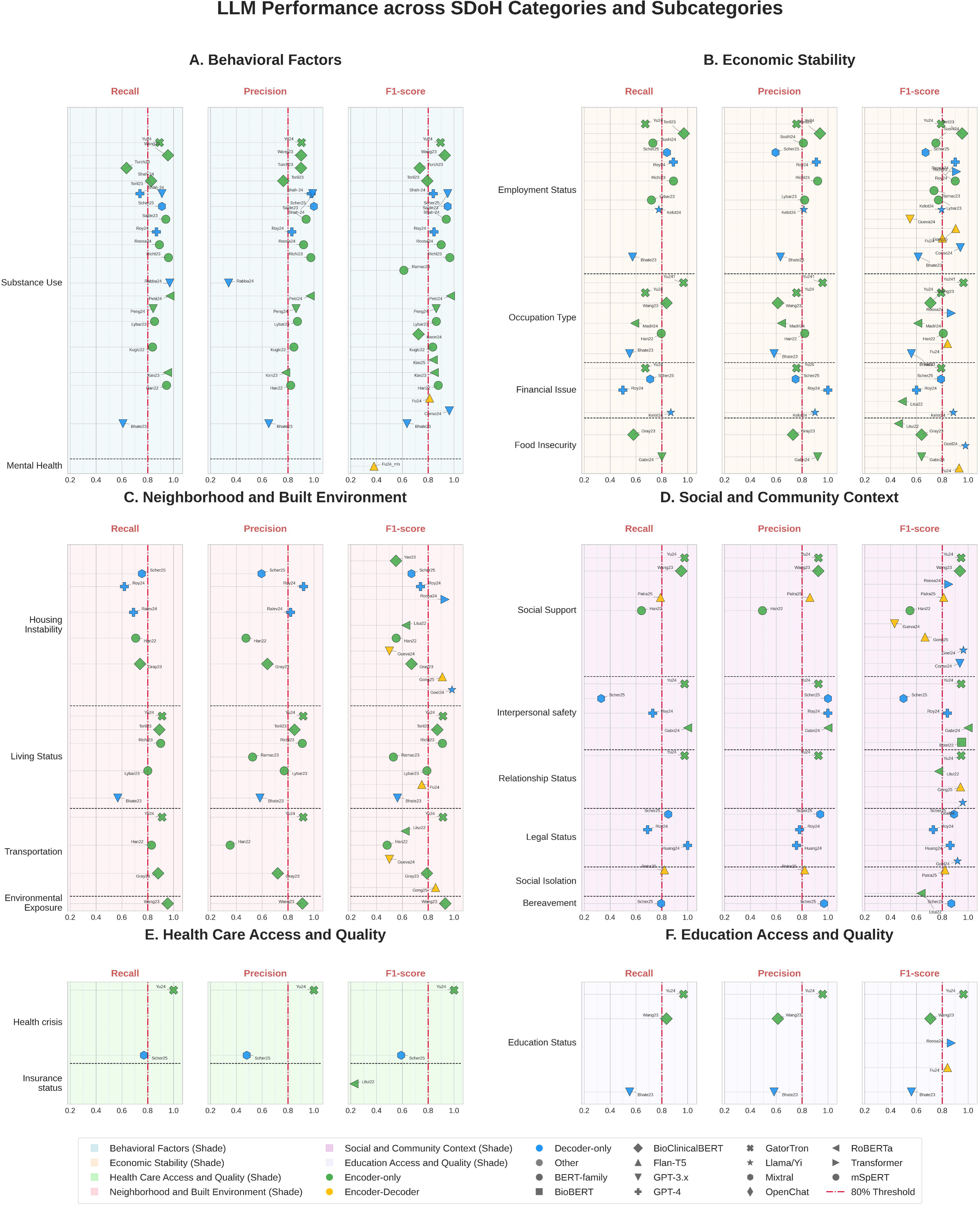
Landscape of LLMs Performance across SDoH Categories and Subcategories. Dot plots display reported performance metrics—Recall, Precision, and F1-score—across studies, grouped by SDoH domain: (A) Behavioral Factors, (B) Economic Stability, (C) Neighborhood and Built Environment, (D) Social and Community Context, (E) Health Care Access and Quality, and (F) Education Access and Quality. Each point represents a metric reported by a single study; point shape and color correspond to the LLM used, as indicated in the in-figure legend. The red dashed line marks the 0.80 performance threshold commonly referenced in clinical benchmarks. Gu (2024) reported only accuracy, and Robitschek (2024) reported F1 < 0.2 across all 27 subcategories. *Abbreviations: LLM, large language model; SDoH, Social Drivers of Health*.

**Behavioral Factors** represented the most consistently high-performing domain (**Figure 3A**). Substance Use extraction achieved outstanding results across several studies, most notably with Petit-Jean 2024’s EDS-CamemBERT (F1=0.972) and Shah-Mohammadi 2024b’s GPT-3.5-turbo (F1=0.95); many models reported F1-scores above the 0.80 clinical benchmark (**Figure 4C**). In contrast, performance in the Mental Health subdomain was significantly lower, with Fu 2024’s Flan-T5-Large (F1=0.38) illustrating the challenges of extracting nuanced psychological concepts.

**Figure 4.**
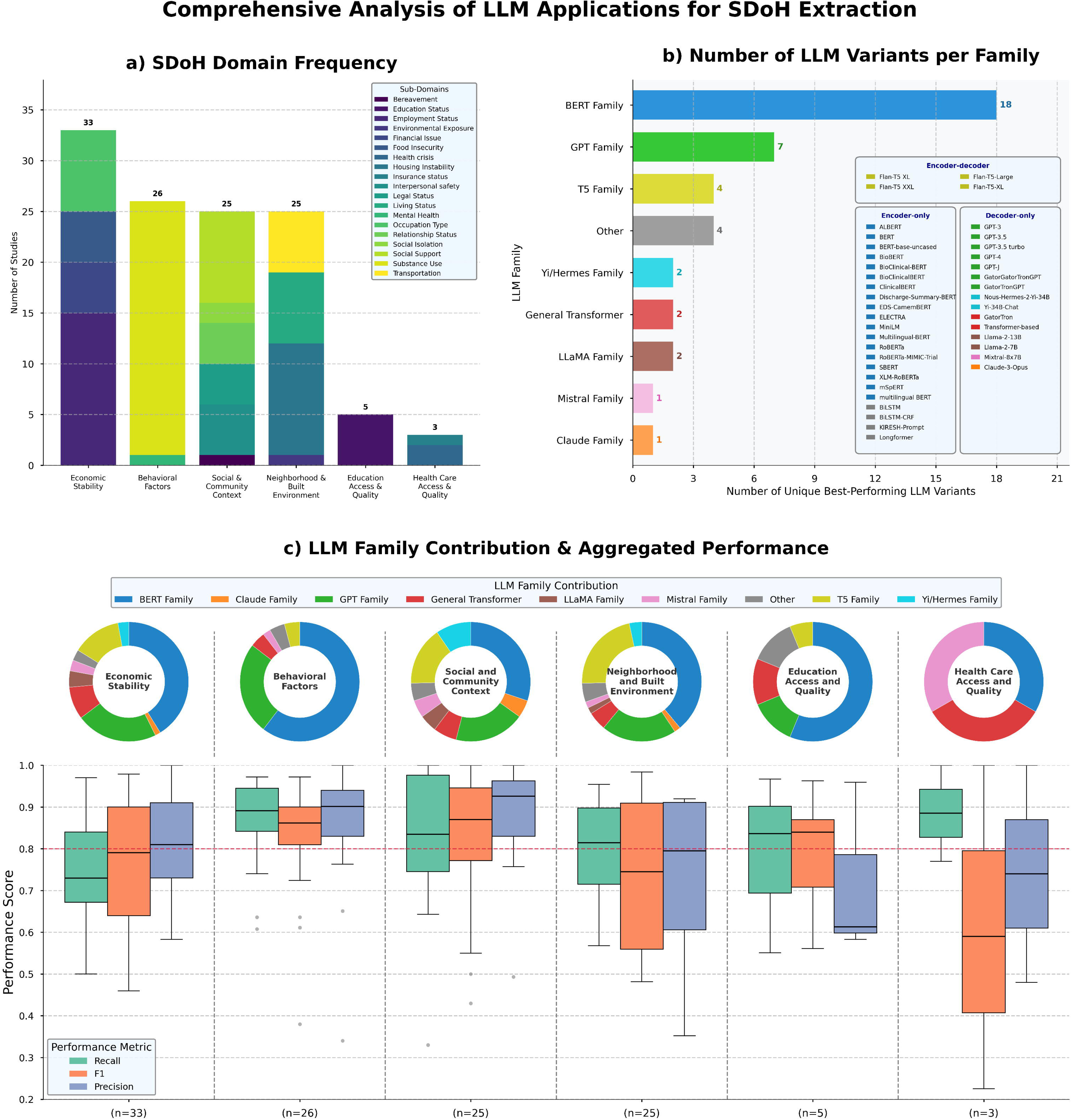
Landscape of LLM Applications in SDoH Extraction. (A) Stacked bar chart showing the number of studies (n = 42) addressing each of the six primary SDoH domains. Bars are color-segmented to represent subdomain-level analysis. (B) Horizontal bar chart displaying the frequency of unique LLM variants used across studies, grouped by model family (e.g., BERT, GPT). (C) Boxplots summarizing Recall, Precision, and F1-score distributions by SDoH domain. Each metric was collected independently, as studies varied in which metrics they reported and which level-2 SDoH subcategories they targeted. As a result, distributions reflect separate collective summaries rather than directly linked values. Boxes represent interquartile ranges (IQR), central lines indicate medians, and whiskers extend to 1.5×IQR. A horizontal red dashed line denotes the 0.80 clinical performance benchmark. Above each domain label, a color-coded donut chart illustrates the proportional frequency of LLM families contributing to the performance data for that domain. *Abbreviations: LLM, large language model; SDoH, Social Drivers of Health*.

**Economic Stability** (**Figure 3 B**; **Figure 4 A**) displayed notable intra-domain variability. Employment Status extraction was among the most successful subdomains, with Consoli 2024’s GPT-3.5 (F1=0.94), Roy 2024’s GPT-4 (F1=0.90), and Torii 2023’s Bio_Discharge_Summary_BERT (F1=0.9538) all demonstrating high precision and recall. In contrast, Financial Issues showed pronounced performance asymmetry, such as Roy 2024’s GPT-4 achieving perfect precision (1.00) but low recall (0.50), resulting in overall moderate F1 (0.60). Food Insecurity displayed the widest variation, with Lituiev 2022’s RoBERTa yielding poor performance (F1=0.46) and Goel 2024’s Yi-34B-Chat achieving near-perfect extraction (F1=0.979).

**Neighborhood and Built Environment** results were mixed (**Figure 3 C**). Living Status frequently achieved strong F1-scores with Yu 2024’s GatorTron (F1=0.914), Richie 2023’s BioClinicalBERT (F1=0.91), and Torii 2023’s Bio_Discharge_Summary_BERT (F1=0.8702), although results ranged from as low as F1=0.53 to above 0.91. Housing Instability showed extreme variability, with F1-values from Han 2022’s BERT (0.552) to Goel 2024’s Nous-Hermes-2-Yi-34B (0.984). Transportation exhibited pronounced precision-recall tradeoffs (e.g., Han 2022: recall=0.829, precision=0.352), raising concerns about false positive rates.

**Social and Community Context** was one of the best-performing and most frequently evaluated domains (**Figure 3 D; Figure 4 A**). Social Support extraction achieved consistent and high F1-scores across multiple models, such as Yu 2024’s GatorTron (F1=0.946), Goel 2024’s Llama-2-13B-chat-hf (F1=0.963), and Wang 2023’s BioClinicalBERT (F1=0.937). Other subdomains, including Relationship Status (F1=0.946–0.958) and Bereavement (F1=0.87), also exhibited reliably strong performance. However, Interpersonal Safety presented greater extraction challenges, often with models achieving high precision (e.g., Roy 2024, Gabriel 2024: precision=1.00) but only moderate recall (Roy 2024: 0.73).

**Health Care Access and Quality** was underrepresented and showed the weakest performance overall (**Figure 3 E; Figure 4 A**). Insurance Status extraction was particularly poor (Lituiev 2022’s RoBERTa: F1=0.225), and Health Crisis detection varied widely, ranging from Yu 2024’s GatorTron (F1=1.00) to Scherbakov 2025’s Mixtral 8×7B (F1=0.59).

**Education Access and Quality** was evaluated in relatively few studies but included examples of strong model performance (**Figure 3 F; Figure 4 A**). Yu 2024’s GatorTron achieved the best results for Education Status (F1=0.963, recall=0.967).

Boxplots in Figure 4 C summarize broader domain-level trends, highlighting Substance Use, Living Status, and Environmental Exposure as high-performing subdomains (median F1 > 0.85), while Mental Health and Insurance Status remain challenging. Figure 4B illustrates the diversity and prevalence of LLM architectures within each domain.

Overall, these results (**Figure 3 and Figure 4**) demonstrate a heterogeneous LLM performance landscape in SDoH extraction. Domains such as behavioral factors and social context yield the most reliable results, while healthcare access and mental health subdomains continue to present distinct challenges.

### 4.3. Methodological Assessment

Our methodological assessment revealed substantial variability in how current studies address internal validity, external validity, and reporting transparency in LLM-based extraction of social determinants of health (**Figure 5**).

**Figure 5.**
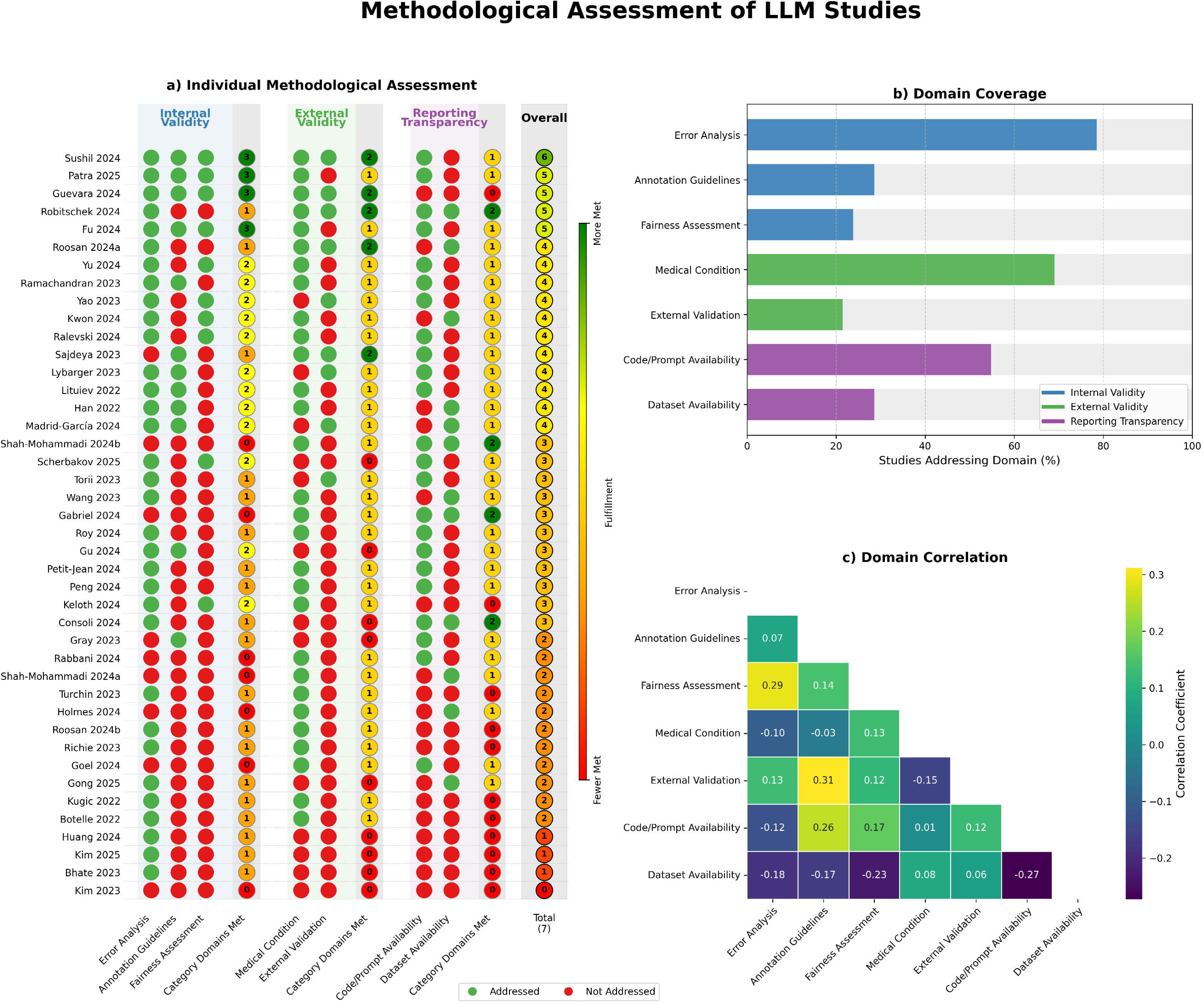
Methodological Evaluation of LLM-Based SDoH Studies. (A) Heatmap showing domain-level methodological assessment for each of the 42 included studies. Green dots indicate the domain was addressed; red dots indicate it was not. (B) Horizontal bar chart summarizing the proportion and count of studies meeting criteria for each of the seven methodological domains, grouped by category: Internal Validity, External Validity, and Reporting Transparency. (C) Correlogram depicting Pearson correlation coefficients between methodological domains, with warmer colors (green) reflecting positive correlations and cooler colors (purple) indicating negative correlations in reporting practices. *Abbreviations: LLM, large language model*

**Internal validity**—which assesses model rigor in identifying limitations and subgroup performance—was partially addressed in most studies. While **error analysis** was the most commonly fulfilled domain (76%, 32/42), **annotation guidelines** were documented in only **29%** (12/42) of studies, and **fairness assessments** across demographic subgroups were reported in just **24%** (10/42). This underreporting limits insight into both label consistency and equity in model behavior—core elements for extracting nuanced and context-dependent SDoH information.

**External validity** was less consistently demonstrated. Although **clinical context or patient population** was specified in **69%** (29/42) of studies, indicating some attention to where models were applied, only **21%** (9/42) conducted external dataset validation using independent datasets— undermining confidence in model generalizability across institutions or care settings.

**Reporting transparency** showed mixed adoption. **Code or prompt availability** was reported in **55%** (23/42) of studies, supporting some reproducibility. However, only **29%** (12/42) made annotated datasets publicly accessible, limiting opportunities for external benchmarking and comparative evaluation—particularly relevant for studies focused on housing instability, employment insecurity, or food access.

Correlational patterns revealed structural tensions and opportunities for synergy. **Dataset availability** was negatively correlated with both **fairness assessment** (r = –0.23) and **error analysis** (r = –0.18), suggesting that privacy concerns or proprietary constraints may hinder both data sharing and rigorous performance evaluation. Conversely, the availability of **annotation guidelines** was moderately associated with **external validation** (r = 0.31), implying that better documentation may support transferability and generalization testing.

A few studies—such as **Sushil 2024** and **Guevara 2024**—met most criteria across internal and external validity dimensions, reflecting high methodological rigor. Yet even among these exemplars, reporting transparency features were not uniformly applied. This inconsistency underscores a broader issue: predictive performance alone is insufficient. Reliable and equitable SDoH extraction depends on comprehensive methodological practices that enhance generalizability, transparency, and fairness.

## 5. DISCUSSION

The influence of SDoH on clinical outcomes is well established, yet health systems continue to face barriers in systematically integrating these factors into routine care.^96–98^ A key limitation is the lack of structured SDoH data—less than 1.5% of content in EHRs is coded, with the majority embedded in free-text narratives.^99^ This review systematically examined how LLMs have been applied to extract SDoH from clinical text and assessed their readiness for clinical deployment using a seven-domain methodological framework that evaluates internal validity, external validity, and reporting transparency.

To our knowledge, this is the first review to systematically evaluate LLMs for SDoH extraction from a methodological perspective. Prior systematic reviews, such as Patra et al., 2021 primarily examined traditional machine learning and rule-based approaches, focusing on a narrow set of SDoH categories and conducted before the emergence of powerful generative models.^100^ More recent scoping reviews, including Li et al., 2024, offered broader overviews of the SDoH data pipeline—from data collection to intervention—but did not appraise LLM-based extraction methods in depth.^101^ Domain-specific reviews, such as those by McNeill et al., 2023 in cardiovascular medicine and Abbott et al., 2024 in emergency care, provided valuable insights yet relied on literature searches completed before key developments in LLM research.^102,103^ Meanwhile, works like Wu et al., 2023 are limited to single-method evaluations of pre-transformer models.^104^ In contrast, our review provides an updated synthesis of LLM performance across domains and introduces a standardized, domain-level evaluation framework tailored to SDoH extraction— accounting for challenges such as linguistic ambiguity, fairness, and contextual nuance.

By integrating WHO- and HHS-endorsed taxonomies with a hierarchical model classification (e.g., GPT, LLaMA, Flan-T5), we enabled structured comparisons across architectures and domains. Our March 2025 search cutoff ensured coverage of recent trends such as open-source LLMs, hybrid prompting strategies, and interpretability methods. Furthermore, current evaluation tools (e.g., HELM, TRIPOD-AI) offer partial guidance but are not optimized for the specific challenges of SDoH extraction.^52,105–107^ Our domain-level framework addresses this gap by providing structured, interpretable evaluation criteria that emphasize transparency, generalizability, and fairness. As such, this work offers the most comprehensive and methodologically rigorous synthesis to date, establishing a foundation for equitable, reliable clinical applications.

LLMs offered substantial advantages over manual annotation methods, which are costly and time-intensive. Ralevski et al. estimated that annotating 25,000 clinical notes cost ∼$9,400 manually compared to <$150 with GPT-3.5.^28^ Consoli et al. similarly showed their SDoH-GPT system to be over 20× cheaper and 10× faster than human annotation, while achieving greater consistency.^94^ These developments paralleled a broader transition from BERT-based models toward encoder-decoder and decoder-only architectures, including privacy-preserving open-source options like LLaMA and Mixtral.^108,109^

Importantly, larger models did not always confer performance advantages. For instance, a fine-tuned LLaMA-2 7B model achieved an F1-score of 0.885 in extracting financial issues— rivaling GPT-4.^94^ Techniques such as domain routing, exemplified by Goel et al.’s Oracle Router, which sends clinical notes to the best-performing model per SDoH domain, further enhanced efficiency and specialization.^93^ However, advanced methods like retrieval-augmented generation and knowledge distillation remain underutilized in this space.^110,111^

Despite these gains, many studies (26%) reported only macro-averaged F1-scores without accompanying recall, precision, or class distribution metrics, limiting interpretability and clinical trust. Low recall may result in missed opportunities for intervention, while low precision could lead to unnecessary screening. Future studies should include class-level metrics, prevalence data, and precision–recall curves to better capture model behavior in real-world scenarios.^112^

Methodological inconsistencies also hindered comparability. Few studies systematically tested prompting strategies; Consoli et al. was an exception, showing variable performance across zero-, two-, and eight-shot prompts.^94^ Additionally, Chain-of-thought prompting and model confidence assessments were rare, with Scherbakov et al.’s study a notable outlier.^113^ They introduced a self-consistency method—accepting outputs only if generated in ≥2 of 4 LLM runs— enhancing reliability without sacrificing efficiency.^59^ Although open-source models are increasingly used, few studies conducted head-to-head comparisons with high-performing proprietary LLMs such as Gemini and Claude. Including these models in future evaluations could enhance generalizability and better reflect real-world clinical utility.^114,115^

Across the three dimensions of internal validity, external validity, and reporting transparency, we observed substantial and recurrent gaps that undermine the reproducibility, equity, and clinical applicability of LLM-based SDoH extraction. Only four studies satisfied all internal validity criteria, and just 29% reported annotation guidelines—highlighting the urgent need for tailored reporting standards specific to this domain. While open science practices are gradually improving, with 55% of studies sharing code or prompts, dataset sharing remains limited (29%), likely constrained by privacy regulations. Interestingly, We observed that studies sharing datasets were less likely to include fairness assessments (r = –0.23) or detailed error analyses (r = –0.18). This inverse association may reflect the tension between data transparency and privacy-preserving documentation practices, where concerns about re-identification or institutional review constraints limit the granularity of performance reporting.

Although the majority of studies included some form of error analysis (86%), only 24% incorporated fairness assessments, and fewer still performed subgroup analyses. These omissions are especially concerning in the context of SDoH, where model predictions may reflect—and potentially reinforce—existing disparities tied to race, gender, socioeconomic status, or insurance type.^29–32,53,54^ Without systematic equity evaluations, such models risk perpetuating structural inequities under the guise of automation.

External validation was also limited. While 69% of studies stated the medical condition evaluated, only 21% tested external datasets, and just four met both criteria. The heavy reliance on private, U.S.-based ICU datasets further limits generalizability. A model trained on discharge notes from urban ICUs may not transfer to outpatient or community settings.^44,116^ This challenge is compounded by the lack of high-quality, publicly available SDoH datasets. Existing corpora are narrow in scope and often rely on surface-level named entity recognition, which fails to capture contextually rich and implicit social information.^117^ Without granular annotations, models lack semantic depth and struggle with nuanced understanding, limiting cross-setting applicability.^118,119^ Additionally, the absence of standardized SDoH taxonomies impedes precision. Most studies used broad domains or basic NER tags that fail to capture conceptually distinct yet clinically relevant distinctions (e.g., eviction risk vs. homelessness).^118,119^ ICD-10 Z-codes, while useful, are too coarse for nuanced modeling.^120^ This lack of shared vocabulary affects interoperability and intervention targeting.^39,121^

To address these challenges, future work should prioritize: (1) standardizing annotation and fairness reporting practices; (2) promoting secure data-sharing infrastructures that preserve patient privacy; and (3) investing in diverse, real-world benchmark datasets spanning care settings, patient populations, and healthcare systems. These efforts are essential for developing trustworthy, generalizable SDoH tools that can be responsibly integrated into clinical workflows.

This review has several limitations. First, the substantial heterogeneity across studies—in annotation scope, modeling strategies, and outcome definitions—precluded the use of formal meta-analytic techniques. To address this, we applied a structured qualitative synthesis supported by a standardized classification framework. Second, due to inconsistent reporting of class distributions, we relied on macro-averaged F1-scores, which may obscure model performance on rare SDoH subcategories. To mitigate this, we provide detailed subdomain-level results in the supplementary materials. Third, for consistency in evaluation, we extracted the best-performing model from each study, which may underrepresent within-study performance variability. Fourth, our search was restricted to English-language studies published prior to March 2025, though we included five major biomedical and technical databases to maximize coverage. Finally, while our methodological assessment framework was designed to be practical and domain-relevant, it remains preliminary and unvalidated. It does not yet weight criteria based on clinical impact—an important area for future refinement, particularly with respect to fairness and generalizability.

## 6. CONCLUSION

LLMs showed strong promise for scalable SDoH extraction from clinical text, but progress is hindered by methodological shortcomings and inconsistent practices. Performance disparities across domains highlight the need for improved rigor, transparency, and contextual sensitivity. Advancing clinically meaningful and generalizable systems will require not just technical innovation but a shift in research culture.

## Supporting information

PRISMA-ScR

## 7. DECLARATIONS

**Funding**: This research received no funding from any source.

**Ethics approval**: Not applicable.

**Consent to participate:** Not applicable.

**Consent for publication:** Not applicable.

**Data availability:** Data sharing is not applicable to this article as no datasets were generated or analyzed during the current study.

**Competing interests:** The authors declare no competing interests.

## Data Availability

All data produced in the present work are contained in the manuscript

## Acknowledgments

None.

## Author Contributions

**M.R.** conceptualized the study. **A.F.,** and **A.G.** designed the methodology. **A.F.** and **A.S**. conducted the data collection and formal analysis. **M.R., A.G.,** and **E.H**. Supervision, Writing – Review & Editing. **A.F.** wrote the original draft and created the visualizations. All authors reviewed and edited the final manuscript

