## Supplementary Table for "Beyond Metrics to Methods: A Scoping Review of Large Language Models for Detection of Social Drivers of Health in Clinical Notes"

**Supplementary Table**. General studies’ characteristics

| **Study** | **Country** | **Dataset ^(a)^** | **Availability** | **Notes** | **Medical Condition(s) &/or Populations** | **LLM(s)^(b)^*** |
| --- | --- | --- | --- | --- | --- | --- |
| Patra 2025 ^56^ | USA | MSHS + WCM | Private | 525 | Psychiatry | **FLAN-T5-XL**, GPT 4.0 ^(c)^, BERT |
| Kim 2025 ^57^ | Korea | SNUH + KBSMC | Private | 5313 | Not specified | **XLM-RoBERTa, Multilingual BERT** |
| Scherbakov 2025 ^58^ | USA | MUSC | Private | 430 | Not specified | Mixtral 8×7B model |
| Rabbani 2024 ^59^ | USA | Stanford Children’s Health | Private | 300 | Adolescent | GPT-3.5 |
| Shah-Mohammadi 2024a ^60^ | USA | MIMIC-III | Public | 100 | Critical care | GPT-4 |
| Gu 2024 ^61^ | USA | MGB system | Private | 200 | Not specified | **openchat 3.5,** zephyr-7b-beta, vicuna-7b-v1.5, Llama-2-7b-chat-hf, vicuna-13b-v1.5, WizardLM-13B-v1.2, Llama-2-13b-chat-hf |
| Shah-Mohammadi 2024b ^62^ | USA | MIMIC-III | Public | 500 | Critical care with a history of COPD | GPT-3.5-turbo |
| Huang 2024 ^63^ | USA | Emergency Departments | Private | 1000 | Not specified | RoBERTa, **GPT-4,** Longformer |
| Roosan 2024a ^64^ | USA | MIMIC | Public | 100 | Critical care | Transformer-based |
| Fu 2024 ^65^ | USA | UW hospital system | Private | 1,260 | Pediatrics | mSpERT (BioClinicalBERT), **Flan-T5-Large**, GPT-4 |
| Guevara 2024 ^66^ | USA | MGB system | Private | 1200 | Oncology | **GPT-3.5**^(c)^**,** Bert-base-uncased, Flan-T5 base large, Flan-T5 L, **Flan-T5 XL,** Flan-T5 XXL |
| Madrid-GarcÃ­a 2024 ^67^ | Spain | MEDDOPROF | Public | 2000 | Not specified | RoBERTa - es |
| Yu 2024 ^68^ | USA | UF Health- IDR | Private | 829 | Cancer | GatorTron |
| Peng 2024 ^69^ | USA | SHAC | Private | 4480 | Critical care + Inpatient | GatorTronGPT (GPT-3) |
| Sushil 2024 ^70^ | USA | SHAC | Private | 500 | Critical care + Inpatient | BERT |
| Keloth 2024 ^71^ | USA | HCPC, UTP, MIMIC-III, Mayo-clinic | Private | 4027 | Psychiatry, Critical care, Chronic pain | SBERT, **LLAMA 2 7B** |
| Kwon 2024 ^72^ | USA | MIMIC-IV | Public | 750 | Critical care + Inpatient | BioBERT, **BioClinicalBERT** |
| Holmes 2024 ^73^ | USA | MIMIC-IV | Public | 50,000 | Critical care | Biomed-Roberta-snli-multinli-stsb |
| Roosan 2024b ^74^ | USA | Emanate Health System | Private | 890 | OUD with a history of Diabetes & Hypertension | BERT |
| Petit-Jean 2024 ^75^ | France | AP-HP | Private | 300 | Cardiology, Oncology, Rheumatology | EDS-CamemBERT |
| Roy 2024 ^27^ | USA | MIMIC-III | Public | 621 | Critical care | GPT-3, **GPT-4** |
| Gabriel 2024 ^76^ | USA | i2b25, MIMIC-III | Public | 1384 | Critical care + Cardiovascular diseases | GPT 3.5 turbo^(c)^, BERT, **RoBERTa** |
| Robitschek 2024 ^77^ | USA | UCSF | Private | 101 | Liver transplant | GPT-4-Turbo-128k |
| Ralevski 2024 ^28^ | USA | Providence Health and Services | Private | 539 | Pregnant women | **GPT-4,** GPT-3.5 turbo |
| Yao 2023 ^78^ | USA | VHA - CDW | Private | 5,000 | Not specified | KiRESH-Prompt (BioClinicalBERT) |
| Ramachandran 2023 ^79^ | USA | SHAC | Private | 4405 | Critical care + Inpatient | GPT-4, **mSpERT** |
| Turchin 2023 ^80^ | USA | MGB EHRs | Private | 3882 | Cardiovascular | BERT, **ClinicalBERT** |
| Wang 2023 ^81^ | USA | MIMIC-SBDH | Public | 7,025 | Critical care | BiLSTM-CRF, BERT, ALBERT, BioBERT, **BioClinicalBERT**, ELECTRA, RoBERTa, RoBERTa-MIMIC-Trial, BiLSTM |
| Richie 2023 ^82^ | USA | SHAC | Private | 3529 | Critical care + Inpatient | BioClinicalBERT |
| Bhate 2023 ^83^ | USA | IUPUI | Private | 1,000 | Not specified | **GPT-3.5,** all-MiniLM-L6-v2 |
| Kim 2023 ^84^ | Korea | SNUH | Private | 4,996 | Not specified | **XLM-RoBERTa**, Multilingual-BERT |
| Gray 2023 ^85^ | USA | JHHS | Private | 192 | Not specified | ClinicalBERT |
| Sajdeya 2023 ^86^ | USA | UF-Shands | Private | 463 | Preoperative patients | **BERT,** BioClinicalBERT |
| Lituiev 2022 ^87^ | USA | UCSF | Private | 1576 | Chronic Pain (Lower Back) | RoBERTa |
| Kugic 2022 ^88^ | Austria | DBM4PM | Private | 1429 | Cardiology, Oncology, Dermatology | German BERT |
| Botelle 2022 ^89^ | UK | CRIS | Private | 3771 | Psychiatry | BioBERT |
| Han 2022 ^26^ | USA | MIMIC-III | Public | 3504 | Critical care | BERT |
| Lybarger 2023 ^90^ | USA | SHAC | Private | 4405 | Critical care + Inpatient | mSpERT |
| Gong 2025 ^91^ | USA | MIMIC-III | Public | 840 | Critical care | BERT, FLAN-T5-Large, **FLAN-T5-XL,** GPT turbo-0301^(c)^ |
| Goel 2024 ^92^ | USA | MIMIC-III | Public | 500 | Critical care | **Claude-3-opus**^(c)^, **Oracle Router^(d)^:** Nous-Hermes-2, Yi-34B, Yi-34B-Chat, Llama-2-13b-chat-hf |
| Consoli 2024 ^93^ | USA | MIMIC-III | Public | 4332 | Critical care | GPT-3.5 |
| Torii 2023 ^94^ | USA | SHAC | Private | 1877 | Critical care + Inpatient | **Bio_Discharge_Summary_BERT,** GPT-J |
| **AP-HP**: Assistance Publique–Hôpitaux de Paris; **BERT**: Bidirectional Encoder Representations from Transformers; **CDW**: Corporate Data Warehouse; **COPD**: Chronic Obstructive Pulmonary Disease; **CRIS**: Clinical Record Interactive Search; **DBM4PM**: Digital Biomarkers for Precision Medicine; **EHR**: Electronic Health Record(s); **GPT**: Generative Pre-trained Transformer; **HCPC**: UTHealth Harris County Psychiatric Center; **HCSC-MSKC**: Hospital Clinico San Carlos Musculoskeletal Cohort; **IDR**: Integrated Data Repository; **i2b2**: Informatics for Integrating Biology and the Bedside; **IUPUI**: Indiana University–Purdue University Indianapolis; **JHHS**: Johns Hopkins Health System; **KBSMC**: Kangbuk Samsung Medical Center; **LLM**: Large Language Model; **Mayo**: Mayo Clinic and the Olmsted Medical Center; **MEDDOPROF**: Medical Occupation Annotation Corpus; **MGB**: Mass General Brigham; **MIMIC**: Medical Information Mart for Intensive Care; **Mistral**: A foundational language model developed by Mistral AI; **MSHS**: Mount Sinai Health System; **MUSC**: Medical University of South Carolina; **OUD**: Opioid Use Disorder; **SBDH**: Social and Behavioral Determinants of Health; **SHAC**: Social History Annotation Corpus; **SNUH**: Seoul National University Hospital; **T5**: Text-to-Text Transfer Transformer; **UF**: University of Florida; **UCSF**: University of California, San Francisco; **UCSD**: University of California, San Diego; **UK**: United Kingdom; **USA**: United States of America; **UTP**: UTHealth Physicians; **UW**: University of Washington; **WCM**: Weill Cornell Medicine.  *For studies evaluating multiple LLMs, the best-performing model(s) are highlighted in bold.  (a) Detailed descriptions of the datasets used in the included studies can be found in **Supplementary Material (Appendix B, Section 2).**  (b) Comprehensive information on the LLMs reviewed is available in **Supplementary Material (Appendix B, Section 3).**  (c) The LLM is used either exclusively for synthetic data generation or in combination with classification tasks.  (d) The Oracle Router dynamically selects the top-performing LLM based on the specific SDoH category: **Nous-Hermes-2-Yi-34B** for homelessness, incarceration, and marital estrangement; **Yi-34B-Chat** for food insecurity; and **Llama-2-13b-chat-hf** for Relative Needing Care. | | | | | | |
