## Supplementary material for "Beyond Metrics to Methods: A Scoping Review of Large Language Models for Detection of Social Drivers of Health in Clinical Notes": PRISMA-ScR

**Preferred Reporting Items for Systematic reviews and Meta-Analyses extension for Scoping Reviews (PRISMA-ScR) Checklist**

| **SECTION** | **ITEM** | **PRISMA-ScR CHECKLIST ITEM** | **REPORTED ON PAGE #** |
| --- | --- | --- | --- |
| **TITLE** | | | |
| Title | 1 | Identify the report as a scoping review. | **Title page (p.1):** "Beyond Metrics to Methods: A Scoping Review of LLMs for SDoH Detection in Clinical Notes" |
| **ABSTRACT** | | | |
| Structured summary | 2 | Provide a structured summary that includes (as applicable): background, objectives, eligibility criteria, sources of evidence, charting methods, results, and conclusions that relate to the review questions and objectives. | **Abstract (p.2):** Provides a structured summary including background, objectives, methods, results, and conclusions. |
| **INTRODUCTION** | | | |
| Rationale | 3 | Describe the rationale for the review in the context of what is already known. Explain why the review questions/objectives lend themselves to a scoping review approach. | **Introduction (p.3-5):** Details the importance of SDoH, challenges with EHR data, the potential of LLMs, and the fragmented nature of current research, justifying comprehensive mapping. |
| Objectives | 4 | Provide an explicit statement of the questions and objectives being addressed with reference to their key elements (e.g., population or participants, concepts, and context) or other relevant key elements used to conceptualize the review questions and/or objectives. | **Introduction (p.5):** Explicitly states three objectives: (1) map SDoH domains and LLM architectures, (2) compare model performance, and (3) assess methodological rigor. |
| **METHODS** | | | |
| Protocol and registration | 5 | Indicate whether a review protocol exists; state if and where it can be accessed (e.g., a Web address); and if available, provide registration information, including the registration number. | **Methods (p.5):** States the review follows PRISMA-ScR and ENTREQ guidelines. No protocol registration is mentioned. |
| Eligibility criteria | 6 | Specify characteristics of the sources of evidence used as eligibility criteria (e.g., years considered, language, and publication status), and provide a rationale. | **Methods (p.6-7):** Provides detailed inclusion and exclusion criteria, specifying study design, use of LLMs, SDoH focus, clinical text data, and required performance metrics. |
| Information sources* | 7 | Describe all information sources in the search (e.g., databases with dates of coverage and contact with authors to identify additional sources), as well as the date the most recent search was executed. | **Methods (p.6):** Lists five databases (PubMed/MEDLINE, Web of Science, Embase, Scopus, IEEE Xplore). Search conducted up to March 1, 2025. |
| Search | 8 | Present the full electronic search strategy for at least 1 database, including any limits used, such that it could be repeated. | **Methods (p.6):** Refers to Appendix B–Section 1 for the full electronic search strategies for all databases. |
| Selection of sources of evidence† | 9 | State the process for selecting sources of evidence (i.e., screening and eligibility) included in the scoping review. | **Methods (p.7):** Details the screening process using Covidence, with calibration, two independent reviewers (A.F. and A.S.), and conflict resolution. |
| Data charting process‡ | 10 | Describe the methods of charting data from the included sources of evidence (e.g., calibrated forms or forms that have been tested by the team before their use, and whether data charting was done independently or in duplicate) and any processes for obtaining and confirming data from investigators. | **Methods (p.7-8):** Describes the use of a standardized extraction form, piloted on 10 studies, with independent data extraction by two reviewers and validation by a third. |
| Data items | 11 | List and define all variables for which data were sought and any assumptions and simplifications made. | **Methods (p.8):** Lists five categories of extracted data: (1) Study Characteristics, (2) Model Information, (3) Dataset Characteristics, (4) SDoH Categories, and (5) LLM Performance. |
| Critical appraisal of individual sources of evidence§ | 12 | If done, provide a rationale for conducting a critical appraisal of included sources of evidence; describe the methods used and how this information was used in any data synthesis (if appropriate). | **Methods (p.11-13):** Details the rationale for and development of a structured methodological assessment framework across seven domains to evaluate research rigor, tailored to SDoH extraction challenges. |
| Synthesis of results | 13 | Describe the methods of handling and summarizing the data that were charted. | **Methods (p.8-11):** Explains the hierarchical classification for SDoH and LLMs, the use of macro-averaging, and the calculation of summary statistics for performance metrics. |
| **RESULTS** | | | |
| Selection of sources of evidence | 14 | Give numbers of sources of evidence screened, assessed for eligibility, and included in the review, with reasons for exclusions at each stage, ideally using a flow diagram. | **Results (p.13) and Figure 2 (p.14):** PRISMA flow diagram shows 254 records from databases plus 18 from snowballing, leading to 42 included studies. |
| Characteristics of sources of evidence | 15 | For each source of evidence, present characteristics for which data were charted and provide the citations. | **Results (p.14) and Table 2 (p.15-17):** Presents detailed characteristics for all 42 included studies. |
| Critical appraisal within sources of evidence | 16 | If done, present data on critical appraisal of included sources of evidence (see item 12). | **Results (p.23-26) and Figures 7-9:** Presents the results of the methodological assessment across all studies. |
| Results of individual sources of evidence | 17 | For each included source of evidence, present the relevant data that were charted that relate to the review questions and objectives. | **Results (p.18-23), Figure 3, and Figure 5:** Presents detailed performance results across SDoH subcategories and aggregated domain-level performance. |
| Synthesis of results | 18 | Summarize and/or present the charting results as they relate to the review questions and objectives. | **Results (p.13-26):** Provides a comprehensive synthesis of findings regarding data sources, LLM architectures, performance trends, and methodological rigor. |
| **DISCUSSION** | | | |
| Summary of evidence | 19 | Summarize the main results (including an overview of concepts, themes, and types of evidence available), link to the review questions and objectives, and consider the relevance to key groups. | **Discussion (p.26-31):** Provides a detailed summary and interpretation of findings, linking inconsistent performance and methodological gaps back to the review's objectives. |
| Limitations | 20 | Discuss the limitations of the scoping review process. | **Discussion (p.31):** Acknowledges limitations, including study heterogeneity precluding meta-analysis, use of macro-averaged F1-scores, and the preliminary nature of the methodological framework. |
| Conclusions | 21 | Provide a general interpretation of the results with respect to the review questions and objectives, as well as potential implications and/or next steps. | **Discussion (p.26-31) and Conclusion (p.31):** Summarizes the implications for clinical deployment and outlines future research directions to address methodological gaps. |
| **FUNDING** | | | |
| Funding | 22 | Describe sources of funding for the included sources of evidence, as well as sources of funding for the scoping review. Describe the role of the funders of the scoping review. | **Title page (p.1):** "This research received no funding from any source." |

JBI = Joanna Briggs Institute; PRISMA-ScR = Preferred Reporting Items for Systematic reviews and Meta-Analyses extension for Scoping Reviews.

* Where *sources of evidence* (see second footnote) are compiled from, such as bibliographic databases, social media platforms, and Web sites.

† A more inclusive/heterogeneous term used to account for the different types of evidence or data sources (e.g., quantitative and/or qualitative research, expert opinion, and policy documents) that may be eligible in a scoping review as opposed to only studies. This is not to be confused with *information sources* (see first footnote).

‡ The frameworks by Arksey and O’Malley (6) and Levac and colleagues (7) and the JBI guidance (4, 5) refer to the process of data extraction in a scoping review as data charting*.*

§ The process of systematically examining research evidence to assess its validity, results, and relevance before using it to inform a decision. This term is used for items 12 and 19 instead of "risk of bias" (which is more applicable to systematic reviews of interventions) to include and acknowledge the various sources of evidence that may be used in a scoping review (e.g., quantitative and/or qualitative research, expert opinion, and policy document).

*From:* Tricco AC, Lillie E, Zarin W, O'Brien KK, Colquhoun H, Levac D, et al. PRISMA Extension for Scoping Reviews (PRISMAScR): Checklist and Explanation. Ann Intern Med. 2018;169:467–473. [doi: 10.7326/M18-0850](http://annals.org/aim/fullarticle/2700389/prisma-extension-scoping-reviews-prisma-scr-checklist-explanation).
