## Appendix A for "Beyond Metrics to Methods: A Scoping Review of Large Language Models for Detection of Social Drivers of Health in Clinical Notes"

**ENTREQ Statement**

| Section | Component | Details |
| --- | --- | --- |
| 1. Aim | Research objective | To comprehensively evaluate Large Language Model (LLM) applications for extracting Social Drivers of Health (SDoH) from clinical text by pursuing three objectives:   1. Systematically map the SDoH domains and LLM architectures investigated to identify research gaps. 2. Quantitatively compare model performance across SDoH categories using standardized metrics to establish benchmarks. 3. Rigorously assess methodological rigor across domains covering internal validity, external validity, and reporting transparency. |
| 2. Synthesis Methodology | Framework and approach | - Followed PRISMA-ScR (Preferred Reporting Items for Systematic Reviews and Meta-Analyses Extension for Scoping Reviews) and ENTREQ (Enhancing Transparency in Reporting the Synthesis of Qualitative Research) guidelines. - Developed and applied a novel methodological framework to standardize SDoH/LLM classification, synthesize performance, and conduct a structured assessment of research rigor. - Utilized a standardized data extraction process and systematic reporting of performance metrics (F1-score, precision, recall). |
| 3. Approach To Searching | Search strategy | - A comprehensive search was conducted in five electronic databases: PubMed/MEDLINE, Web of Science Core Collection, Embase, Scopus, and IEEE Xplore. - Reference lists of relevant reviews were manually screened to identify additional studies ("snowballing"). - Search strategies combined controlled vocabulary (e.g., MeSH) and free-text keywords. - Queries were structured around three core concepts: (1) Large Language Models, (2) Social Drivers of Health, and (3) Clinical Text Data. |
| 4. Inclusion Criteria | Eligibility criteria | **Inclusion:**   - Clinical notes, EHR free-text, or synthetic clinical narratives focusing on SDOH - Large language models for SDOH extraction tasks - Quantitative performance metrics - Original research with empirical evaluation   **Exclusion:**   - Traditional ML approaches without TLM comparison - Non-clinical text - Secondary research - Lacking comprehensive evaluation metrics |
| 5. Data Sources | Search parameters | - Time period: January 1, 2018, to March 1, 2025 - Language: English - Document types: Peer-reviewed journals, conference proceedings, high-quality preprints - Initial search: January 15, 2025 - Update: March 1, 2025 |
| 6. Electronic Search Strategy | Search terms | - Transformer language models: "GPT," "BERT," "transformer architecture" - Social determinants of health: "health equity," "food insecurity," "housing instability" - Clinical text data: "EHR free-text," "clinical narratives" - Complete strategies documented in supplementary materials (**Appendix B**) |
| 7. Study Screening Methods | Selection process | - Deduplication using Mendeley (v1.19.8) - Pilot screening of 10 citations (Cohen's kappa > 0.8) - Independent title/abstract screening by two reviewers (A.F. and A.S.) - Full-text assessment - Disagreement resolution through discussion - Documented using PRISMA 2020 flow diagram |
| 8. Study Characteristics | Overview of included studies | - 42 studies total. - Geographic distribution: United States (36), South Korea (2), Spain (1), France (1), Austria (1), UK (1). - Datasets: Private institutional (29 studies, 69%), publicly available (13 studies, 31%). - Sample sizes: Ranged from 100 to 50,000 notes. - Models: Diverse architectures including BERT variants, GPT variants (GPT-3 to GPT-4), T5 variants (Flan-T5), LLaMA, and Mixtral. - Most studied specialty: Critical care (19 studies, 45.2%), largely due to the use of MIMIC datasets. |
| 9. Study Selection Results | PRISMA flow | - Initial records from databases: 254. - Records after duplicates removed: 159. - Additional records from "snowballing": 18. - Total records for screening: 177. - Excluded after title/abstract screening: 45. - Full-text articles assessed for eligibility: 132. - Articles excluded after full-text review: 90. - Final included studies: 42. - Documented in PRISMA flow diagram (Figure 2). |
| 10. Rationale For Appraisal | Quality assessment framework | A structured methodological assessment framework was developed to address the unique challenges of SDoH extraction (e.g., ambiguous concepts, sparse documentation, equity concerns).  The framework is organized across three dimensions: Internal Validity, External Validity, and Reporting Transparency.  It is presented as a pragmatic, exploratory tool for evaluating research rigor, not a validated scoring instrument.  Full rationale and criteria are available in the manuscript (Section 2.6) and **Appendix C.** |
| 11. Appraisal Items | Quality assessment focus | - **Internal Validity:** Error Analysis, Annotation Guidelines, Fairness Assessment. - **External Validity:** External Dataset Validation, Medical Condition Specificity. - **Reporting Transparency:** Code/Prompt Availability, Dataset Availability. |
| 12. Appraisal Process | Quality assessment methods | Two reviewers independently assessed each of the 42 studies against the seven domains of the methodological framework. The domains cover internal validity, external validity, and reporting transparency. |
| 13. Appraisal Results | Quality assessment findings | - Error Analysis was the most common practice (79% of studies). - Critical gaps were identified: only 29% provided Annotation Guidelines, 24% conducted Fairness Assessment, and 21% performed External Dataset Validation. - Overreliance on MIMIC datasets from U.S. critical care settings (12 studies). - Four studies (Guevara 2024, Fu 2024, Sushil 2024, Patra 2025) met all three internal validity domains. - Limited availability of public, annotated datasets hinders reproducibility and benchmarking. |
| 14. Data Extraction | Extraction methodology | **Extraction methodology:**   - A standardized extraction form was piloted on 10 studies. - Two independent reviewers with complementary expertise: A.F. (pharmacist, health informatics) and A.S. (computer scientist, clinical NLP). - A third reviewer (M.R.) validated 20% of extractions (Cohen's kappa = 0.92).   **Data extracted:**   - Study Characteristics (author, year, country, setting). - Model Information (architecture, parameters, fine-tuning). - Dataset Characteristics (source, size, availability, annotation). - SDoH Categories (domains, subdomains, frameworks). - LLM Performance (precision, recall, F1-score, etc.). |
| 15. Software | Tools used | - Citation management: Mendeley (v1.19.8) - Screening: Covidence (Veritas Health Innovation) - Data management: Excel (v2502) - Analysis/visualization: Excel (v2502), Python 3.10 (Matplotlib, Seaborn) |
| 16. Number Of Reviewers | Reviewer roles | - Two independent reviewers (A.F. and A.S.) for screening and data extraction. - Third reviewer (M.R.) for validation of extracted data. - All reviewers for resolving disagreements to reach consensus. |
| 17. Coding | SDOH classification | **Framework**: Primarily based on the U.S. HHS Healthy People 2030 framework, supplemented by WHO and CDC domains.  **Six domains:**   - Economic Stability - Education Access and Quality - Health Care Access and Quality - Neighborhood and Built Environment - Social and Community Context - Behavioral Factors (included as a distinct category for this review's specific aims).   Performance metrics were aggregated to the Level-2 (subcategory) granularity. |
| 18. Study Comparison | Comparative analysis | - **Metrics**: Recall, precision, F1-score. - For studies with multiple models, the single best-performing model was selected based on the highest F1-score, and all associated metrics (recall, precision, F1) for that specific model were extracted. - Lenient matching metrics were prioritized for span extraction tasks to better capture clinical utility. - Summary statistics (min, Q1, median, Q3, max) were calculated for each SDoH domain. |
| 19. Derivation Of Themes | Key themes identified | 1. Significant variability in LLM performance across SDoH domains. 2. Widespread methodological gaps, particularly in fairness, external validation, and transparency. 3. Heavy reliance on a limited number of datasets (e.g., MIMIC) and a lack of geographic/clinical diversity, threatening generalizability. 4. An evolving architectural landscape moving beyond BERT to more diverse and open-source models. 5. The absence of standardized SDoH classification schemes and benchmark datasets, which impedes progress. 6. The need for more nuanced evaluation beyond the F1-score to assess clinical and equitable utility. |
| 20. Quotations | Use of quotations | Not applicable (quantitative synthesis of performance metrics and methodological practices). |
| 21. Synthesis Output | Key findings and implications | **Performance variation (Median F1-score):**   - **Strong**: Behavioral Factors (Median F1: 0.87). - **Moderate**: Social and Community Context (Median F1: 0.86), Neighborhood and Built Environment (Median F1: 0.77). - **Challenging**: Economic Stability (characterized by low recall, median: 0.72), Education Access & Quality (characterized by low precision, median: 0.61). - **Poor/Volatile**: Health Care Access & Quality (Median F1: 0.59).   **Future directions:**   - Adopt standardized SDoH classification schemes. - Prioritize external validation on diverse, representative datasets. - Conduct rigorous fairness audits and subgroup performance analysis. - Develop and share high-quality, context-rich benchmark datasets. - Explore advanced techniques like chain-of-thought prompting and intra-model consistency checks. |
