## Appendix B for "Beyond Metrics to Methods: A Scoping Review of Large Language Models for Detection of Social Drivers of Health in Clinical Notes"

**Section (1)**

**Detailed Search Strategy**

1. **Search Databases**

Five databases were searched: PubMed/MEDLINE, Web of Science, Embase, Scopus, and IEEE Xplore.

2. **Search Queries**
   2.1. **PubMed**

("large language model*"[Title/Abstract] OR "llm"[Title/Abstract] OR "gpt"[Title/Abstract] OR "generative pre trained transformer"[Title/Abstract] OR "transformer*"[Title/Abstract] OR "retrieval augmented generation"[Title/Abstract] OR "prompt engineering"[Title/Abstract] OR "instruction tuning"[Title/Abstract] OR "fine-tuning"[Title/Abstract] OR "one-shot"[Title/Abstract] OR "zero-shot"[Title/Abstract] OR "few-shot"[Title/Abstract]) **AND** ("social determinants of health"[Title/Abstract] OR "sdoh"[Title/Abstract] OR "social behavior*"[Title/Abstract] OR "health disparities"[Title/Abstract] OR "health equity"[Title/Abstract] OR "social risk factors"[Title/Abstract] OR "social needs"[Title/Abstract] OR "healthy people 2030"[Title/Abstract] OR "income inequality"[Title/Abstract] OR "unemployment"[Title/Abstract] OR "poverty"[Title/Abstract] OR "food insecurity"[Title/Abstract] OR "housing instability"[Title/Abstract] OR "economic stability"[Title/Abstract] OR "financial stress"[Title/Abstract] OR "homelessness"[Title/Abstract] OR "education disparity"[Title/Abstract] OR "health literacy"[Title/Abstract] OR "access to healthcare"[Title/Abstract] OR "transportation access"[Title/Abstract] OR "healthcare disparities"[Title/Abstract] OR "neighborhood safety"[Title/Abstract] OR "environmental hazards"[Title/Abstract] OR "discrimination"[Title/Abstract] OR "violence"[Title/Abstract] OR "substance use"[Title/Abstract] OR "alcohol"[Title/Abstract] OR "tobacco"[Title/Abstract] OR "drug use"[Title/Abstract] OR "interpersonal violence"[Title/Abstract] OR "social inequities"[Title/Abstract]) **AND** ("electronic health record*"[Title/Abstract] OR "ehr"[Title/Abstract] OR "clinical text"[Title/Abstract] OR "free text"[Title/Abstract] OR "medical notes"[Title/Abstract] OR "clinical narrative"[Title/Abstract] OR "unstructured data"[Title/Abstract])

2.2. **Web of Sciences**

(TI=("large language model*" OR "llm" OR "transformer*" OR "gpt" OR "generative pre trained transformer" OR "retrieval augmented generation" OR "prompt engineering" OR "instruction tuning" OR "fine-tuning" OR "one-shot" OR "zero-shot" OR "few-shot") OR AB=("large language model*" OR "llm" OR "gpt" OR "generative pre trained transformer" OR "retrieval augmented generation" OR "prompt engineering" OR "instruction tuning" OR "fine-tuning" OR "one-shot" OR "zero-shot" OR "few-shot" OR "transformer*")) **AND** (TI=("social determinants of health" OR "sdof" OR "social behavior*" OR "health disparities" OR "health equity" OR "social risk factors" OR "social needs" OR "healthy people 2030" OR "income inequality" OR "unemployment" OR "poverty" OR "food insecurity" OR "housing instability" OR "economic stability" OR "financial stress" OR "homelessness" OR "education disparity" OR "health literacy" OR "access to healthcare" OR "transportation access" OR "healthcare disparities" OR "neighborhood safety" OR "environmental hazards" OR "discrimination" OR "violence" OR "substance use" OR "alcohol" OR "tobacco" OR "drug use" OR "interpersonal violence" OR "social inequities") OR AB=("social determinants of health" OR "sdof" OR "social behavior*" OR "health disparities" OR "health equity" OR "social risk factors" OR "social needs" OR "healthy people 2030" OR "income inequality" OR "unemployment" OR "poverty" OR "food insecurity" OR "housing instability" OR "economic stability" OR "financial stress" OR "homelessness" OR "education disparity" OR "health literacy" OR "access to healthcare" OR "transportation access" OR "healthcare disparities" OR "neighborhood safety" OR "environmental hazards" OR "discrimination" OR "violence" OR "substance use" OR "alcohol" OR "tobacco" OR "drug use" OR "interpersonal violence" OR "social inequities")) **AND** (TI=("electronic health record*" OR "ehr" OR "clinical text" OR "free text" OR "medical notes" OR "clinical narrative" OR "unstructured data") OR AB=("electronic health record*" OR "ehr" OR "clinical text" OR "free text" OR "medical notes" OR "clinical narrative" OR "unstructured data"))

2.3. **Embase**

('large language model*':ti,ab,kw OR 'llm':ti,ab,kw OR 'gpt':ti,ab,kw OR 'generative pre-trained transformer':ti,ab,kw OR 'transformer*':ti,ab,kw OR 'retrieval augmented generation':ti,ab,kw OR 'prompt engineering':ti,ab,kw OR 'instruction tuning':ti,ab,kw OR 'fine-tuning':ti,ab,kw OR 'one-shot':ti,ab,kw OR 'zero-shot':ti,ab,kw OR 'few-shot':ti,ab,kw) **AND** ('social determinants of health'/exp OR 'social determinants of health':ti,ab,kw OR 'sdoh':ti,ab,kw OR 'social behavior*':ti,ab,kw OR 'health disparities'/exp OR 'health disparities':ti,ab,kw OR 'health equity'/exp OR 'health equity':ti,ab,kw OR 'social risk factors' OR 'social risk factors':ti,ab,kw OR 'social needs':ti,ab,kw OR 'healthy people 2030':ti,ab,kw OR 'income inequality'/exp OR 'income inequality':ti,ab,kw OR 'unemployment':ti,ab,kw OR 'poverty':ti,ab,kw OR 'food insecurity'/exp OR 'food insecurity':ti,ab,kw OR 'housing instability'/exp OR 'housing instability':ti,ab,kw OR 'economic stability'/exp OR 'economic stability':ti,ab,kw OR 'financial stress':ti,ab,kw OR 'homelessness':ti,ab,kw OR 'education disparity' OR 'education disparity':ti,ab,kw OR 'health literacy'/exp OR 'health literacy':ti,ab,kw OR 'access to healthcare'/exp OR 'access to healthcare':ti,ab,kw OR 'transportation access' OR 'transportation access':ti,ab,kw OR 'healthcare disparities'/exp OR 'healthcare disparities':ti,ab,kw OR 'neighborhood safety'/exp OR 'neighborhood safety':ti,ab,kw OR 'environmental hazards' OR 'environmental hazards':ti,ab,kw OR 'discrimination':ti,ab,kw OR 'violence':ti,ab,kw OR 'substance use'/exp OR 'substance use':ti,ab,kw OR 'alcohol':ti,ab,kw OR 'tobacco':ti,ab,kw OR 'drug use'/exp OR 'drug use':ti,ab,kw OR 'interpersonal violence'/exp OR 'interpersonal violence':ti,ab,kw OR 'social inequities' OR 'social inequities':ti,ab,kw) **AND** ('electronic health record'/exp OR 'electronic health record*':ti,ab,kw OR 'ehr':ti,ab,kw OR 'clinical text':ti,ab,kw OR 'free text':ti,ab,kw OR 'medical notes':ti,ab,kw OR 'clinical narrative':ti,ab,kw OR 'unstructured data':ti,ab,kw)

2.4. **Scopus**

TITLE-ABS-KEY (( "large language model*" OR llm OR gpt OR "generative pre-trained transformer" OR "retrieval augmented generation" OR "prompt engineering" OR "instruction tuning" OR "fine-tuning" OR "one-shot" OR "zero-shot" OR "few-shot" OR "*transformer*" ) **AND** ( "social determinants of health" OR sdoh OR "social behavior*" OR "health disparities" OR "health equity" OR "social risk factors" OR "social needs" OR "healthy people 2030" OR "income inequality" OR unemployment OR poverty OR "food insecurity" OR "housing instability" OR "economic stability" OR "financial stress" OR homelessness OR "education disparity" OR "health literacy" OR "access to healthcare" OR "transportation access" OR "healthcare disparities" OR "neighborhood safety" OR "environmental hazards" OR discrimination OR violence OR "substance use" OR alcohol OR tobacco OR "drug use" OR "interpersonal violence" OR "social inequities" ) **AND** ("electronic health record*" OR ehr OR "clinical text" OR "free text" OR "medical notes" OR "clinical narrative" OR "unstructured data" ))

2.5. **IEEE**

(("large language model*" OR "llm" OR "gpt" OR "generative pre-trained transformer" OR "transformer*" OR "retrieval augmented generation" OR "prompt engineering" OR "instruction tuning" OR "fine-tuning" OR "one-shot" OR "zero-shot" OR "few-shot") **AND** ("social determinants of health" OR "sdoh" OR "social behavior*" OR "health disparities" OR "health equity" OR "social risk factors" OR "social needs" OR "healthy people 2030" OR "income inequality" OR "unemployment" OR "poverty" OR "food insecurity" OR "housing instability" OR "economic stability" OR "financial stress" OR "homelessness" OR "education disparity" OR "health literacy" OR "access to healthcare" OR "transportation access" OR "healthcare disparities" OR "neighborhood safety" OR "environmental hazards" OR "discrimination" OR "violence" OR "substance use" OR "alcohol" OR "tobacco" OR "drug use" OR "interpersonal violence" OR "social inequities") **AND** ("electronic health record*" OR "ehr" OR "clinical text" OR "free text" OR "medical notes" OR "clinical narrative" OR "unstructured data"))

3. **Search Filters:**
   - **Study Type:**

- Prioritize empirical studies (e.g., validation, implementation, or evaluation of LLMs).
- Exclude reviews, editorials, and theoretical papers.
- **Language:** English only (most LLM/SDoH research is published in English).
- **Document Type:** Focus on full-text articles, preprints (e.g., arXiv), and conference papers (for IEEE).

4. **Software and Analytical Tools**

Mendeley (v1.19.8) was used as a citation manager , Covidence (Veritas Health Innovation) ^2^ for screening, Excel (v2502) for data management, and Excel (v2502) along with Python 3.10 (utilizing Matplotlib and Seaborn libraries) for visualization.

**Section (2)**

**Description of Datasets Used in the Studies**

| Dataset | Description | Population/Setting |
| --- | --- | --- |
| MIMIC | De-identified discharge summaries from hospital and emergency department visits. | Beth Israel Deaconess Medical Center, USA |
| MEDDOPROF | Spanish clinical case report corpus with annotations for occupations. | 1,844 Spanish clinical case reports |
| HCSC-MSKC | Cohort with >117,000 visits and >35,000 subjects in rheumatology outpatient clinic (2007–2017). | Hospital Clinico San Carlos, Madrid, Spain |
| SHAC | De-identified clinical text (social history notes) from MIMIC-III and UW, annotated for Social Determinants of Health. | MIMIC-III (USA), University of Washington, USA |
| AP-HP | Multi-hospital EHR dataset comprising 38 university hospitals in Greater Paris area. | Paris, France; 22,000+ beds, 1.5M hospitalizations/year |
| Providence Health Services | Integrated health care system data across urban and rural settings in 7 US states. | USA: AK, CA, MT, OR, NM, TX, WA |
| i2b2/n2c2 | Clinical note datasets for tasks such as deidentification and heart disease cohort selection. | Multiple US institutions |
| UTP | Clinical datasets from UTHealth Physicians. | University of Texas Health, Houston, USA |
| Mayo | Clinical data from Mayo Clinic and Olmsted Medical Center. | Rochester, Minnesota, USA |
| HCPC | Psychiatric clinical records. | UTHealth Harris County Psychiatric Center, Houston, USA |
| IDR | Integrated clinical data repository. | UF Health, Gainesville, Florida, USA |
| MGB | Clinical data from Mass General Brigham healthcare system. | Massachusetts, USA |
| MUSC | Clinical records from the Medical University of South Carolina. | South Carolina, USA |
| SNUH | Family medicine clinic data. | Seoul National University Hospital, South Korea |
| KBSMC | Clinical data from Angbuk Samsung Hospital. | South Korea |
| MSHS | Clinical records from Mount Sinai Health System. | New York, USA |
| WCM | EHR data from Weill Cornell Medicine. | New York, USA |
| CRIS | Clinical Records Interactive Search system. | UK National Health Service hospitals |
| UCSF | Clinical data from University of California, San Francisco. | California, USA |
| IUPUI | Health records from Indiana University–Purdue University Indianapolis. | Indiana, USA |
| DBM4PM | Digital Biomarkers for Precision Medicine project data. | Not specified |
| UCSD | Clinical records from University of California, San Diego. | California, USA |
| JHHS | Clinical and health-related data, including electronic health records (EHRs), research datasets, and population health analytics data from the Johns Hopkins Health System. | Baltimore, Maryland, USA |
| UF-Shands | Clinical data, primarily electronic health records (EHRs) from the UF Health system, including de-identified patient demographics, encounters, diagnoses, procedures, medications, lab results, and clinical notes. | Gainesville, Florida, USA |

**Section (3)**

**Overview of the Large Language Models Employed in the Studies**

**Overview**

This section outlines major language models and variants relevant to SDoH research. It summarizes model families, key versions, developers, licenses, release years, and distinctive features.

| Family | Model | Developer | License | Year | Description / Key Characteristics |
| --- | --- | --- | --- | --- | --- |
| BERT | BERT-Base Uncased | Google | Apache 2.0 | 2018 | 12-layer Transformer encoder, 768 hidden units, 12 attention heads, 110M parameters; uncased (treats lowercase and uppercase the same), trained with Masked Language Modeling (MLM) and Next Sentence Prediction (NSP) on English BookCorpus and Wikipedia. Bidirectional context understanding. |
|  | RoBERTa | Facebook AI (Meta) | MIT | 2019 | Optimized BERT: no NSP, larger dataset |
|  | ALBERT | Google + Toyota Technological Institute | Apache 2.0 | 2019 | Parameter-efficient (layer sharing, SOP) |
|  | BioBERT | Korea University + Clova AI | Apache 2.0 | 2019 | BERT for biomedical texts (PubMed/PMC) |
|  | ClinicalBERT | MIT + Beth Israel Deaconess Medical Center | Apache 2.0 | 2019 | Fine-tuned for clinical notes/EHR |
|  | DistilBERT | Hugging Face | Apache 2.0 | 2019 | Distilled version (smaller, faster) |
|  | XLM-RoBERTa | Facebook AI (Meta) | MIT | 2020 | Multilingual (100+ languages), no NSP |
|  | Multilingual BERT | Google | Apache 2.0 | 2018 | Trained on 104 languages (Wikipedia) |
|  | German-BERT | deepset GmbH | Apache 2.0 | 2019 | German-language BERT |
|  | mSPERT | German Research Center for Artificial Intelligence (DFKI) | MIT | 2021 | Biomedical + clinical data; focuses on biomedical entity linking and relation extraction |
|  | GatorTron | University of Florida (UF Health) | Research-use | 2022 | Large clinical BERT for EHR/NLP |
|  | Bio_ClinicalBERT-KIRESH | KIRESH Lab (AIIMS Rishikesh) | Custom (research) | 2023 | Hybrid of BioBERT + ClinicalBERT |
| GPT | GPT-3 | OpenAI | Proprietary | 2020 | 175B parameters, autoregressive |
|  | GPT-3.5 | OpenAI | Proprietary | 2022 | Improved GPT-3 with RLHF |
|  | GPT-3.5-turbo | OpenAI | Proprietary | 2023 | Optimized for ChatGPT |
|  | GatorTronGPT | University of Florida | Research-use | 2022 | Clinical GPT-3 variant |
|  | GPT-4 | OpenAI | Proprietary | 2023 | Multimodal (text + image), stronger than GPT-3 |
|  | GPT-4-Turbo | OpenAI | Proprietary | 2023 | 128k context window, cost-efficient |
|  | GPT-3.5-Turbo (0301) | OpenAI | Proprietary | 2023 | 16k context window; highly optimized for low-latency and cost-efficiency. |
| LLaMA | Llama-2-7b | Meta | Custom (commercial use allowed) | 2023 | Base 7B parameter model |
|  | Llama-2-13b | Meta | Custom | 2023 | 13B parameter model |
|  | Llama-2-7b-chat | Meta | Custom | 2023 | Chat-optimized 7B model |
|  | Llama-2-13b-chat | Meta | Custom | 2023 | Chat-optimized 13B model |
|  | Vicuna-7B/13B/33B | LMSYS (UC Berkeley, CMU, Stanford) | Non-commercial | 2023 | Fine-tuned LLaMA for chat (v1.3–v1.5) |
|  | Llama-2-13b-chat-hf | Meta / Hugging Face | Custom (Meta) | 2023 | 13B-parameter chat-optimized version of LLaMA 2; available via Hugging Face with use restrictions. |
| T5 | Flan-T5 (Base/Large/XL/XXL) | Google Research | Apache 2.0 | 2022 | Instruction-tuned T5 variants (XXL: 11B) |
| Mixture-of-Experts | Mixtral 8x7B | Mistral AI | Apache 2.0 | 2023 | Sparse MoE (8 experts, 7B each) |
|  | Switch Transformer | Google Research | Apache 2.0 | 2021 | Early MoE model with dynamic routing |
| Instruction-Tuned | WizardLM-13B | NL2AI Lab (Peking University) | Apache 2.0 | 2023 | Fine-tuned for complex instructions |
|  | Zephyr-7B | Hugging Face | Apache 2.0 | 2023 | RLHF-tuned for chat |
|  | OpenChat 3.5 | OpenChat community | Custom | 2023 | GPT-3.5-based conversational model |
| Embedding Models | all-MiniLM-L6-v2 | Microsoft Research | Apache 2.0 | 2021 | Lightweight sentence embeddings |
|  | Sentence-BERT | UKPLab | Apache 2.0 | 2019 | Siamese BERT for sentence similarity |
| Yi | Yi-34B-Chat | 01.AI | Apache 2.0 | 2023 | Chat-tuned variant of Yi-34B; designed for dialogue and interactive applications. |
|  | Yi-34B | 01.AI | Apache 2.0 | 2023 | 34B-parameter base model; multilingual, pre-trained on large-scale diverse corpora. |
| Claude | Claude-3-opus | Anthropic | Proprietary | 2024 | Flagship Claude 3 model, strong performance across reasoning and vision tasks; ~200k context window. |
| Nous | Nous-Hermes-2 | Nous Research | Apache 2.0 | 2024 | Fine-tuned Mistral-7B, optimized for chat and instruction-following tasks. |
| Specialized | Longformer | AllenAI | Apache 2.0 | 2020 | Handles long documents (16k+ tokens) |
|  | ELECTRA | Google Research | Apache 2.0 | 2020 | Efficient (replaced MLM with discriminator) |

- **RoBERTa:** Removed Next Sentence Prediction (NSP) and trained on 160GB of text (vs. BERT’s 16GB).
- **ALBERT:** Uses Sentence Order Prediction (SOP) instead of NSP and cross-layer parameter sharing.
- **ClinicalBERT:** Trained on MIMIC-III EHR dataset.
- **mSPERT:** Focuses on biomedical entity linking and relation extraction.
- **GatorTron/GatorTronGPT:** Available via UF Health with restrictions (not fully open-source).
- **Bio_ClinicalBERT-KIRESH:** License varies by derivative use (contact lab for details).
- **GPT-3.5**: Uses Reinforcement Learning from Human Feedback (RLHF) for alignment.
- **GPT-4 Multimodal:** Image input capability is restricted in public APIs (text-only for most users).
- **LLaMA** **2 License**: Custom Meta license allows commercial use with restrictions (e.g., products with >700 million monthly active users require approval)
- **Vicuna**: Derived from LLaMA; non-commercial due to original LLaMA license terms.
- **Proprietary Models (**OpenAI): Access requires API or partnership.
- **Research-use Models:** Contact developers for access (e.g., GatorTron).
- **License Compliance:** Verify terms for commercial use (e.g., LLaMA 2, Vicuna).

**Section (4)**

**Links for Annotations Guidelines, Code Repositories and Prompt Specifications**

| Authors (Year) | Focus Area | Code | Prompts | Annotation Guidelines |
| --- | --- | --- | --- | --- |
| Patra (2025) | Social support/isolation | [GitHub](https://github.com/CornellMHILab/Social_Support_Social_Isolation_Extraction) | - | [Full-text](https://arxiv.org/pdf/2403.17199) |
| Scherbakov (2025) | Stressful life events | - | [Supplementary](https://static-content.springer.com/esm/art%3A10.1186%2Fs12889-024-21123-2/MediaObjects/12889_2024_21123_MOESM1_ESM.docx) | - |
| Rabbani (2024) | Confidential content | - | [Few-shot Prompt](https://cdn.jamanetwork.com/ama/content_public/journal/peds/939329/pld230063supp1_prod_1708531182.46147.pdf) | - |
| Gu (2024) | SDoH extraction | - | [Prompts](https://www.medrxiv.org/content/medrxiv/early/2024/08/10/2024.08.08.24311237/DC1/embed/media-1.docx?download=true) | [Supplementary](https://www.medrxiv.org/content/medrxiv/early/2024/08/10/2024.08.08.24311237/DC1/embed/media-1.docx?download=true) |
| Shah-Mohammadi (2024) | Substance use | - | [Zero-shot & Few-shot Prompts](https://jmir.org/api/download?alt_name=medinform_v12i1e56243_app1.png) | - |
| Fu (2024) | Pediatric SDoH | [GitHub](https://github.com/uw-bionlp/PedSHAC) | - | [GitHub](https://github.com/uw-bionlp/PedSHAC) |
| Guevara (2024) | SDoH identification | [GitHub](https://github.com/AIM-Harvard/SDoH) | - | [GitHub](https://github.com/AIM-Harvard/SDoH) |
| Madrid-García (2024) | Occupation extraction | - | - | [Supplementary](https://www.medrxiv.org/content/medrxiv/early/2024/05/09/2024.05.08.24306389/DC4/embed/media-4.pdf?download=true) and [Zenodo](https://zenodo.org/records/4720833) |
| Yu (2024) | SDoH extraction | [GitHub](https://github.com/uf-hobi-informatics-lab/SODA_Docker) | - | - |
| Sushil (2024) | SDoH extraction | [GitHub](https://github.com/tuur/sdoh_n2c2track2_ucsf_umcu) and [Code Status](https://github.com/tuur/code-status-annotations-mimic) | - | [GitHub](https://github.com/tuur/sdoh_n2c2track2_ucsf_umcu) |
| Roy (2024) | SBDH detection | - | [Supplementary](https://static-content.springer.com/esm/art%3A10.1186%2Fs12911-024-02705-x/MediaObjects/12911_2024_2705_MOESM1_ESM.pdf) | - |
| Gabriel (2024) | SDoH identification | [GitHub](https://github.com/UCSDGabrielLab/SDoHLL) | [Supplementary](https://www.pnas.org/doi/suppl/10.1073/pnas.2320716121/suppl_file/pnas.2320716121.sapp.pdf) | - |
| Ralevski (2024) | Housing instability in pregnancy | - | [CoT Prompt](https://jmir.org/api/download?alt_name=jmir_v26i1e63445_app1.docx) | - |
| Yao (2023) | Eviction status | [GitHub](https://github.com/seasonyao/KIRESH-Prompt-) | - | [Supplementary](https://oup.silverchair-cdn.com/oup/backfile/Content_public/Journal/jamia/30/8/10.1093_jamia_ocad081/1/ocad081_supplementary_data.pdf) |
| Ramachandran (2023) | SHAC annotation | - | [One-shot Prompt](https://aclanthology.org/2023.clinicalnlp-1.41.pdf) | - |
| Lituiev (2022) | SDoH extraction | [GitHub](https://github.com/BCHSI/social-deternimants-of-health-clbp) | - | [Supplementary](https://oup.silverchair-cdn.com/oup/backfile/Content_public/Journal/jamia/30/8/10.1093_jamia_ocad054/1/ocad054_supplementary_data.zip) |
| Botelle (2022) | Interpersonal violence | - | - | [Supplementary](https://bmjopen.bmj.com/content/bmjopen/12/2/e052911/DC2/embed/inline-supplementary-material-2.pdf?download=true) |
| Han (2022) | SDOH classification | - | - | [Supplementary](https://ars.els-cdn.com/content/image/1-s2.0-S1532046421003130-mmc1.pdf) |
| Gray (2023) | SDOH classification | [GitHub](https://github.com/ggray15jh/sdoh_pipeline.git.) | - | [Supplementary](https://oup.silverchair-cdn.com/oup/backfile/Content_public/Journal/jamiaopen/6/4/10.1093_jamiaopen_ooad085/1/ooad085_supplementary_data.docx?Expires=1750551704&Signature=kAU6UK1HC19v0cL84zgMdxt85k-smCledgfpJpuZlVQLPTZwyC-MThW6IA6UfMx5G-rd5Bi2cNg98-5tvpEGr7Lqirm9v2D6kIu9Q~1rXAv1w3JjYX47-ccFIrATBDuUVEReXFXk60Ebu~4pAhZOnL2JzL6NKVzAnux5wN1P7iT~rs86KEVTgArftrX~Q1tqlq-1ifr~JcFbnUnbWANbnXDRqlCNVrmbkr3atVRWKp-r-aSsZjGbHmdQo5GpU10NBPNsdgbBmQv6D0Ur5Fa-nvuX-~yVjlmosUMXxQ46A8EuF3poUnDlQC6XlJjgnNyimADy~oPyCT-v8jl~syNsNA__&Key-Pair-Id=APKAIE5G5CRDK6RD3PGA) |
| Sajdeya (2023) | SDOH classification | [GitHub](https://github.com/masoud-r/cannabis-use-NLP.) | - | [Supplementary](https://oup.silverchair-cdn.com/oup/backfile/Content_public/Journal/jamia/30/8/10.1093_jamia_ocad080/1/ocad080_supplementary_data.zip?Expires=1750235299&Signature=NXIpojDgo5yT266GKPh67kq~El~WIgyLjSaQIvljK-~vLz24LB70WBbzaLV8JxxKUZil0TqQNC6~Cu-aRSvimqXDvNj5FJED2Diigce1SfUb5pouL1EIxxH-RouRT-nSGemxiDDHoIvsSUW01dgt72vN8QX9SViCPRB4HbR-islfyQQINRrRa2POBKmnmJFFcU9fFyRI2mwmpsYy1GqvQzga4aCUhEm6NdeIBukOsHcenwsTqgoqMK46XR4Aj9Orz-z1yDNnCoTKjIPiW1~IQfuf9jJJ-FJiQy-WpERjTvJktBdplK7Lz~26rOxhdkJilCgYbGa~iWjJ0Y616SPTJA__&Key-Pair-Id=APKAIE5G5CRDK6RD3PGA) |
| Lybarger (2023) | SDOH classification | [GitHub](https://github.com/Lybarger/sdoh_extraction)) | - | [Supplementary](https://www.sciencedirect.com/science/article/pii/S1532046420302598?via%3Dihub) |
| Gong (2025) | SDOH classification | - | - | - |
| Goel (2024) | SDOH classification | - | [Prompts](https://arxiv.org/pdf/2405.19631) | - |
| Consoli (2024) | SDOH classification | - | [Prompts](https://arxiv.org/pdf/2407.17126) | - |
| Torii (2023) | SDOH classification | - | [Prompts](https://arxiv.org/pdf/2301.11386) | - |

**Section (5)**

**Social Drivers of Health (SDoH) and Behavioral Factors: Model Performance by Domain**

**Table of Contents**

1. Overview
2. Economic Stability
3. Education Access and Quality
4. Health Care Access and Quality
5. Neighborhood and Built Environment
6. Social and Community Context
7. Behavioral Factors
8. References

**Overview**

This table presents performance metrics from various studies that developed models to identify social drivers of health (SDOH) and behavioral factors from clinical text. Performance is measured primarily through sensitivity/recall, F1-score, AUC, and precision/PPV.

**Table 1.** Economic Stability

| Study | Fine-granular SDoH | Recall | F1 | Precision | Others |
| --- | --- | --- | --- | --- | --- |
| Scherbakov 2025 | Employment instability | 0.84 | 0.67 | 0.595 | - |
| Scherbakov 2025 | Financial insecurity | 0.71 | 0.79 | 0.75 | - |
| Gu 2024 | Employment Status | - | - | - | 0.79* |
| Roosan 2024a | Employment Status | - | 0.91 | - | - |
| Fu 2024 | Employment Status | - | 0.80 | - | - |
| Fu 2024 | Food Insecurity | - | 0.93 | - | - |
| Guevara 2024 | Employment Status | - | 0.55 | - | - |
| Madrid-García 2024 | Occupation: Profession | 0.59 | 0.61 | 0.64 | - |
| Yu 2024 | Employment stability | 0.672 | 0.7913 | 0.757 | - |
| Yu 2024 | Financial constraint | 0.672 | 0.7913 | 0.757 | - |
| Yu 2024 | Occupation | 0.672 | 0.7913 | 0.757 | - |
| Sushil 2024 | Employment status | 0.73 | 0.75 | 0.81 | - |
| Keloth 2024 | Financial Issues | 0.870 | 0.885 | 0.900 | - |
| Keloth 2024 | Employment Status | 0.777 | 0.795 | 0.814 | - |
| Roy 2024 | Unemployed | 0.89 | 0.90 | 0.91 | - |
| Roy 2024 | Financial circumstance | 0.50 | 0.60 | 1.00 | - |
| Gabriel 2024 | Food Insecurity | 0.8 | 0.64 | 0.92 | 0.78** |
| Robitschek 2024 | Patient's housing situation | 0.00^ | 0.00^ | 0.00^ | - |
| Robitschek 2024 | Adequate health insurance | 0.00^ | 0.00^ | 0.00^ | - |
| Ramachandran 2023 | Employment Status | - | 0.735 | - | - |
| Wang 2023 | Economics | 0.906 | 0.877 | 0.851 | - |
| Richie 2023 | Employment Status | 0.89 | 0.9 | 0.92 | - |
| Bhate 2023 | Employment Status | 0.574 | 0.613 | 0.632 | - |
| Lybarger 2023 | Employment Status | 0.72 | 0.77 | 0.82 | - |
| Gong 2025 | Employment Status | - | 0.9032 | - | - |
| Consoli 2024 | Employment Status | - | 0.94 | - | - |
| Torii 2023 | Employment Status | 0.9701 | 0.9538 | 0.938 |  |
| Lituiev 2022 | Financial strain | - | 0.4868 | - | - |
| Lituiev 2022 | Food Insecurity | - | 0.4583 | - | - |
| Goel 2024 | Food Insecurity | - | 0.979 | - | - |
| Han 2022 | Occupation | 0.796 | 0.807 | 0.821 | 0.874** |
| Gray 2023 | Food Insecurity | 0.58 | 0.64 | 0.73 | - |

***** Accuracy; **Area under the receiver operating curve

^ The performance metrics for this study are derived from 2×2 contingency tables.

**Table 2.** Education Access and Quality

| Study | Fine-granular SDoH | Recall | F1 | Precision | Others |
| --- | --- | --- | --- | --- | --- |
| Gu 2024 | Education Status | - | - | - | 0.94* |
| Roosan 2024a | Education Status | - | 0.87 | - | - |
| Fu 2024 | Education Status | - | 0.84 | - | - |
| Yu 2024 | Education Status | 0.967 | 0.963 | 0.959 | - |
| Wang 2023 | Education Status | 0.836 | 0.708 | 0.613 | - |
| Bhate 2023 | Education Status | 0.551 | 0.561 | 0.583 | - |
| Robitschek 2024 | English-language interpreter needed | 0.00^ | 0.00^ | 0.00^ | - |
| Robitschek 2024 | Understanding of transplant process | 0.00^ | 0.00^ | 0.00^ | - |
| Robitschek 2024 | Insight into liver disease | 0.00^ | 0.00^ | 0.00^ | - |

***** Accuracy; **Area under the receiver operating curve

^ The performance metrics for this study are derived from 2×2 contingency tables.

**Table 3.** Health Care Access and Quality

| Study | Fine-granular SDoH | Recall | F1 | Precision | Others |
| --- | --- | --- | --- | --- | --- |
| Scherbakov 2025 | Health crisis | 0.77 | 0.59 | 0.48 | - |
| Lituiev 2022 | Insurance status | - | 0.2250 | - | - |
| Robitschek 2024 | Receiving treatment for mental health | 0.00^ | 0.00^ | 0.00^ | - |
| Robitschek 2024 | History of medical non-compliance | 0.00^ | 0.00^ | 0.00^ | - |
| Robitschek 2024 | Dishonesty during evaluation | 0.00^ | 0.00^ | 0.00^ | - |
| Robitschek 2024 | Motivation for transplant | 0.00^ | 0.00^ | 0.00^ | - |
| Robitschek 2024 | Addendum with listing decision | 0.00^ | 0.00^ | 0.00^ | - |
| Robitschek 2024 | Transplant listing status | 0.00^ | 0.00^ | 0.00^ | - |
| Yu 2024 | Language | 1.00 | 1.00 | 1.00 | - |

***** Accuracy; **Area under the receiver operating curve

^ The performance metrics for this study are derived from 2×2 contingency tables.

**Table 4.** Neighborhood and Built Environment

| Study | Fine-granular SDoH | Recall | F1 | Precision | Others |
| --- | --- | --- | --- | --- | --- |
| Scherbakov 2025 | Housing instability | 0.755 | 0.67 | 0.595 | - |
| Roosan 2024a | Housing instability | - | 0.93 | - | - |
| Fu 2024 | Living Arrangement | - | 0.75 | - | - |
| Guevara 2024 | Housing instability | - | 0.5 | - | - |
| Guevara 2024 | Transportation barriers | - | 0.5 | - | - |
| Robitschek 2024 | Transportation barriers | 0.00 | 0.00 | 0.00 | - |
| Lituiev 2022 | Transportation barriers | - | 0.6223 | - | - |
| Han 2022 | Transportation barriers | 0.829 | 0.482 | 0.352 | 0.976 |
| Gray 2023 | Transportation barriers | 0.88 | 0.79 | 0.72 | - |
| Gong 2025 | Transportation barriers | - | 0.8571 | - | - |
| Yu 2024 | Living condition | 0.911 | 0.914 | 0.917 | - |
| Yu 2024 | Transportation | 0.911 | 0.914 | 0.917 | - |
| Roy 2024 | Housing Insecurity | 0.62 | 0.74 | 0.92 | - |
| Gray 2023 | Housing Insecurity | 0.74 | 0.67 | 0.64 | - |
| Gray 2023 | Homelessness | 0.62 | 0.68 | 0.81 | - |
| Gabriel 2024 | Homelessness | 0.8 | 0.72 | 0.9 | 0.78** |
| Ralevski 2024 | Housing Instability: General | 0.781 | - | 0.936 | - |
| Ralevski 2024 | Housing Instability: Stable | 0.39 | - | 0.88 | - |
| Ralevski 2024 | Housing Instability: Current Instability | 0.91 | - | 0.83 | - |
| Ralevski 2024 | Housing Instability: Past Instability | 0.68 | - | 0.63 | - |
| Yao 2023 | Eviction: Presence | - | 0.55 | - | - |
| Yao 2023 | Eviction: Period | - | 0.69 | - | - |
| Ramachandran 2023 | Living Status | - | 0.526 |  | - |
| Wang 2023 | Environment | 0.954 | 0.932 | 0.911 | - |
| Richie 2023 | Living status | 0.90 | 0.91 | 0.91 | - |
| Bhate 2023 | Living status | 0.568 | 0.562 | 0.583 | - |
| Lybarger 2023 | Living status | 0.8 | 0.79 | 0.77 | - |
| Torii 2023 | Living status | 0.8906 | 0.8702 | 0.8507 | - |
| Lituiev 2022 | Housing instability | - | 0.6273 | - | - |
| Han 2022 | Housing instability | 0.707 | 0.552 | 0.474 | 0.903** |
| Gong 2025 | Housing instability | - | 0.9091 | - | - |
| Goel 2024 | Housing instability | - | 0.984 | - | - |

***** Accuracy; **Area under the receiver operating curve

^ The performance metrics for this study are derived from 2×2 contingency tables.

**Table 5.** Social and Community Context

| Study | Fine-granular SDoH | Recall | F1 | Precision | Others |
| --- | --- | --- | --- | --- | --- |
| Patra 2025 | Social Support (SS) | 0.79 | 0.81 | 0.86 | - |
| Patra 2025 | Social Isolation (SI) | 0.82 | 0.82 | 0.82 | - |
| Scherbakov 2025 | Bereavement | 0.795 | 0.87 | 0.97 | - |
| Scherbakov 2025 | Criminal justice involvement | 0.85 | 0.89 | 0.94 | - |
| Scherbakov 2025 | Interpersonal safety | 0.33 | 0.50 | 1.00 | - |
| Huang 2024 | Incarceration | 1.00 | 0.862 | 0.757 | - |
| Roosan 2024a | Social Support | - | 0.85 | - | - |
| Fu 2024 | Adoption | - | 0.84 | - |  |
| Fu 2024 | Mental Health | - | 0.38 | - |  |
| Fu 2024 | Trauma | - | 0.60 | - |  |
| Guevara 2024 | Parent | - | 0.54 | - | - |
| Guevara 2024 | Relationship | - | 0.68 | - | - |
| Guevara 2024 | Social Support | - | 0.43 | - | - |
| Yu 2024 | Gender status | 0.976 | 0.946 | 0.926 | - |
| Yu 2024 | Marital status | 0.976 | 0.946 | 0.926 | - |
| Yu 2024 | Partner | 0.976 | 0.946 | 0.926 | - |
| Yu 2024 | Social cohesion | 0.976 | 0.946 | 0.926 | - |
| Yu 2024 | Abuse | 0.976 | 0.946 | 0.926 | - |
| Roy 2024 | Physical & sexual abuse | 0.73 | 0.84 | 1.00 | - |
| Roy 2024 | Legal circumstances | 0.69 | 0.73 | 0.78 | - |
| Gabriel 2024 | Domestic Violence | 1.00 | 1.00 | 1.00 | 0.83** |
| Robitschek 2024 | Designated caregiver | 0.00^ | 0.00^ | 0.00^ | - |
| Robitschek 2024 | Concerns about AF35 ability | 0.00^ | 0.00^ | 0.00^ | - |
| Robitschek 2024 | Barriers to AF38 ability | 0.00^ | 0.00^ | 0.00^ | - |
| Robitschek 2024 | Backup caregiver | 0.00^ | 0.00^ | 0.00^ | - |
| Robitschek 2024 | Past trauma affecting well-being | 0.00^ | 0.00^ | 0.00^ | - |
| Robitschek 2024 | Overall psychosocial risk | 0.00^ | 0.00^ | 0.00^ | - |
| Robitschek 2024 | Psychosocial recommendation | 0.00^ | 0.00^ | 0.00^ | - |
| Wang 2023 | Community | 0.95 | 0.937 | 0.925 | - |
| Lituiev 2022 | Marital or partnership status | - | 0.7718 | - | - |
| Lituiev 2022 | Social isolation | - | 0.6363 | - | - |
| Botelle 2022 | Interpersonal violence: Presence | - | 0.95 | - | - |
| Botelle 2022 | Interpersonal violence: Perpetrator | - | 0.85 | - | - |
| Botelle 2022 | Interpersonal violence: Victim | - | 0.90 | - | - |
| Botelle 2022 | Interpersonal violence: Domestic | - | 0.93 | - | - |
| Botelle 2022 | Interpersonal violence: Physical | - | 0.98 | - | - |
| Botelle 2022 | Interpersonal violence: Sexual | - | 0.93 | - | - |
| Han 2022 | Social Environment | 0.796 | 0.807 | 0.821 | 0.874** |
| Gong 2025 | Parentship | - | 0.8889 | - | - |
| Gong 2025 | Relationship | - | 0.9928 | - | - |
| Gong 2025 | Social Support | - | 0.6667 | - | - |
| Goel 2024 | Incarceration | - | 0.918 | - | - |
| Goel 2024 | Marital Estrangement | - | 0.958 | - | - |
| Goel 2024 | Relative Needing Care | - | 0.963 | - | - |
| Consoli 2024 | Social Support | - | 0.9357 | - | - |
| Han 2022 | Support Circumstances and Networks | 0.643 | 0.550 | 0.493 | 0.844** |

***** Accuracy; **Area under the receiver operating curve

^ The performance metrics for this study are derived from 2×2 contingency tables.

**Table 6.** Behavioral Factors

| Study | Fine-granular SDoH | Recall | F1 | Precision | Others |
| --- | --- | --- | --- | --- | --- |
| Kim 2025 | Alcohol Consumption Pattern: Current status | - | 0.874 | - | - |
| Kim 2025 | Alcohol Consumption Pattern: Amount | - | 0.80 | - | - |
| Kim 2025 | Alcohol Consumption Pattern: Binge | - | 0.842 | - | - |
| Scherbakov 2025 | Mental health crisis | 0.91 | 0.95 | 1.0 | - |
| Rabbani 2024 | Substance use (+ mental health + sexual health) | 0.97 | - | 0.34 | - |
| Shah-Mohammadi 2024a | Tobacco | 0.75 | 0.85 | 0.97 | - |
| Shah-Mohammadi 2024a | Drug | 0.78 | 0.87 | 0.99 | - |
| Shah-Mohammadi 2024a | Alcohol | 0.69 | 0.81 | 0.99 | - |
| Gu 2024 | Tobacco | - | - | - | 0.96* |
| Gu 2024 | Alcohol | - | - | - | 0.86* |
| Gu 2024 | Illicit drugs | - | - | - | 0.86* |
| Gu 2024 | Exercise | - | - | - | 0.86* |
| Shah-Mohammadi 2024b | Tobacco | 0.93 | 0.96 | 0.98 | - |
| Shah-Mohammadi 2024b | Drug | 0.92 | 0.95 | 0.99 | - |
| Shah-Mohammadi 2024b | Alcohol | 0.89 | 0.94 | 0.99 | - |
| Fu 2024 | Substance Use | - | 0.81 | - | - |
| Yu 2024 | Alcohol_use | 0.891 | 0.895 | 0.905 | - |
| Yu 2024 | Tobacco_use | 0.891 | 0.895 | 0.905 | - |
| Yu 2024 | Drug_use | 0.891 | 0.895 | 0.905 | - |
| Yu 2024 | Physical_activity | 0.891 | 0.895 | 0.905 | - |
| Peng 2024 | Substance use (Alcohol) | 0.8424 | 0.8615 | 0.8615 | - |
| Peng 2024 | Substance use (Drug) | 0.8424 | 0.8615 | 0.8615 | - |
| Peng 2024 | Substance use (Tobacco) | 0.8424 | 0.8615 | 0.8615 | - |
| Peng 2024 | Physical activity | 0.8424 | 0.8615 | 0.8615 | - |
| Kwon 2024 | Confirmed Aberrant Behaviors | - | 0.8736 | - | - |
| Kwon 2024 | Suggested Aberrant Behaviors | - | 0.5730 | - | - |
| Roosan 2024b | Opioid use disorder (OUD) | 0.89 | 0.9 | 0.92 | - |
| Petit-Jean 2024 | Alcohol Consumption | 1.00 | 1.00 | 1.00 | - |
| Petit-Jean 2024 | Tobacco consumption | 0.943 | 0.943 | 0.943 | - |
| Roy 2024 | Tobacco use | 0.93 | 0.90 | 0.88 | - |
| Roy 2024 | Opiate abuse | 0.83 | 0.74 | 0.67 | - |
| Roy 2024 | Alcohol abuse | 0.90 | 0.86 | 0.82 | - |
| Roy 2024 | Cocaine abuse | 0.81 | 0.88 | 0.95 | - |
| Robitschek 2024 | Mental health issues affecting functioning | 0.00^ | 0.00^ | 0.00^ | - |
| Robitschek 2024 | Evidence of alcohol abuse/addiction | 0.00^ | 0.00^ | 0.00^ | - |
| Robitschek 2024 | Severity of past alcohol use | 0.00^ | 0.00^ | 0.00^ | - |
| Robitschek 2024 | Current alcohol use | 0.00^ | 0.00^ | 0.00^ | - |
| Robitschek 2024 | Alcohol use in past 6 months | 0.00^ | 0.00^ | 0.00^ | - |
| Robitschek 2024 | Alcohol use in past year | 0.00^ | 0.00^ | 0.00^ | - |
| Robitschek 2024 | Substance use with health concerns | 0.00^ | 0.00^ | 0.00^ | - |
| Robitschek 2024 | Healthy coping strategies | 0.00^ | 0.00^ | 0.00^ | - |
| Ramachandran 2023 | Alcohol | - | 0.694 | - | - |
| Ramachandran 2023 | Drug | - | 0.426 | - | - |
| Ramachandran 2023 | Tobacco | - | 0.714 | - | - |
| Turchin 2023 | Tobacco use status: Never | 0.691 | 0.785 | 0.963 | - |
| Turchin 2023 | Tobacco use status: Past | 0.640 | 0.728 | 0.876 | - |
| Turchin 2023 | Tobacco use status: Current | 0.577 | 0.688 | 0.865 | - |
| Wang 2023 | Alcohol Use | 0.966 | 0.935 | 0.906 | - |
| Wang 2023 | Tobacco Use | 0.957 | 0.915 | 0.875 | - |
| Wang 2023 | Drug Use | 0.944 | 0.935 | 0.926 | - |
| Richie 2023 | Alcohol | 0.98 | 0.98 | 0.99 | - |
| Richie 2023 | Drug | 0.93 | 0.95 | 0.97 | - |
| Richie 2023 | Tobacco | 0.97 | 0.97 | 0.97 | - |
| Bhate 2023 | Alcohol | 0.614 | 0.652 | 0.683 | - |
| Bhate 2023 | Tobacco | 0.631 | 0.652 | 0.662 | - |
| Bhate 2023 | Drug use | 0.579 | 0.603 | 0.609 | - |
| Kim 2023 | Alcohol Consumption | 0.9533 | 0.847 | 0.7823 | - |
| Kugic 2022 | Alcohol consumption: Current drinker | 0.68 | 0.79 | 0.93 | - |
| Kugic 2022 | Alcohol consumption: Current non-drinker | 0.94 | 0.88 | 0.83 | - |
| Kugic 2022 | Alcohol consumption: Ex-problem drinker | 0.92 | 0.92 | 0.92 | - |
| Kugic 2022 | Alcohol consumption: Disorder caused by alcohol | 0.78 | 0.88 | 1.00 | - |
| Kugic 2022 | Alcohol consumption: Problem drinker | 0.84 | 0.82 | 0.81 | - |
| Kugic 2022 | Alcohol consumption: Unknown | 0.86 | 0.82 | 0.78 | - |
| Han 2022 | Substance Use | 0.945 | 0.877 | 0.821 | 0.984** |
| Lybarger 2023 | Alcohol | 0.84 | 0.86 | 0.88 | - |
| Lybarger 2023 | Drug | 0.87 | 0.87 | 0.87 | - |
| Lybarger 2023 | Tobacco | 0.85 | 0.86 | 0.87 | - |
| Consoli 2024 | Tobacco | - | 0.9631 | - | - |
| Torii 2023 | Alcohol | 0.9273 | 0.8947 | 0.8644 | - |
| Torii 2023 | Drug | 0.6667 | 0.6487 | 0.6316 | - |
| Torii 2023 | Tobacco | 0.8841 | 0.8356 | 0.7922 | - |
| Sajdeya 2023 | Cannabinoids | 0.94 | 0.94 | 0.94 | - |

***** Accuracy; **Area under the receiver operating curve

^ The performance metrics for this study are derived from 2×2 contingency tables.

**Table 7.** Overview of LLM performance for SDoH categories consolidated to level-2 granularity

| SDoH Category | Subcategory | Study | Best performing LLM ^(a)^ | Recall | F1 | Precision |
| --- | --- | --- | --- | --- | --- | --- |
| Economic Stability | **Employment Status** | Scherbakov 2025 ^1^ | Mixtral 8 ×7B model | 0.84 | 0.67 | 0.595 |
|  |  | Roosan 2024a ^2^ | Transformer-based | - | 0.91 | - |
|  |  | Fu 2024 ^3^ | Flan-T5-Large | - | 0.80 | - |
|  |  | Guevara 2024 ^4^ | Flan-T5 XL + GPT-3.5 ^(b)^ | - | 0.55 | - |
|  |  | Yu 2024 ^5^ | GatorTron | 0.672 | 0.791 | 0.757 |
|  |  | Sushil 2024 ^6^ | BERT | 0.73 | 0.75 | 0.81 |
|  |  | Keloth 2024 ^7^ | LLAMA 2 7B | 0.777 | 0.795 | 0.814 |
|  |  | Roy 2024 ^8^ | GPT-4 | 0.89 | 0.90 | 0.91 |
|  |  | Ramachandran 2023 ^9^ | mSpERT | - | 0.735 | - |
|  |  | Richie 2023 ^10^ | BioClinical-BERT | 0.89 | 0.9 | 0.92 |
|  |  | Lybarger 2023 ^11^ | mSpERT | 0.72 | 0.77 | 0.82 |
|  |  | Gong 2025 ^12^ | FLAN-T5-XL + GPT turbo-0301 | - | 0.9032 | - |
|  |  | Consoli 2024 ^13^ | GPT-3.5 | - | 0.94 | - |
|  |  | Torii 2023 ^14^ | Bio_Discharge_Summary_BERT | 0.9701 | 0.9538 | 0.938 |
|  |  | Bhate 2023 ^15^ | GPT-3.5 | 0.574 | 0.613 | 0.632 |
|  | **Financial Issue** | Scherbakov 2025 ^1^ | Mixtral 8 ×7B model | 0.71 | 0.79 | 0.75 |
|  |  | Yu 2024 ^5^ | GatorTron | 0.672 | 0.791 | 0.757 |
|  |  | Keloth 2024 ^7^ | LLAMA 2 7B | 0.870 | 0.885 | 0.900 |
|  |  | Roy 2024 ^8^ | GPT-4 | 0.50 | 0.60 | 1.00 |
|  |  | Lituiev 2022 ^16^ | RoBERTa | - | 0.49 | - |
|  | **Food Insecurity** | Fu 2024 ^3^ | Flan-T5-Large | - | 0.93 | - |
|  |  | Gabriel 2024 ^17^ | RoBERTa + GPT-3.5 turbo ^(b)^ | 0.80 | 0.64 | 0.92 |
|  |  | Lituiev 2022 ^16^ | RoBERTa | - | 0.46 | - |
|  |  | Gray 2023 ^18^ | ClinicalBERT | 0.58 | 0.64 | 0.73 |
|  |  | Goel 2024 ^19^ | Yi-34B-Chat | - | 0.979 | - |
|  | **Occupation Type** | Madrid-García 2024 ^20^ | RoBERTa - es | 0.59 | 0.61 | 0.64 |
|  |  | Yu 2024 ^5^ | GatorTron | 0.672 | 0.791 | 0.757 |
|  |  | Han 2022 ^21^ | BERT | 0.796 | 0.807 | 0.821 |
|  |  | Roosan 2024a ^2^ | Transformer-based | - | 0.87 | - |
|  |  | Fu 2024 ^3^ | Flan-T5-Large | - | 0.84 | - |
|  |  | Yu 2024 ^5^ | GatorTron | 0.967 | 0.963 | 0.959 |
|  |  | Wang 2023 ^22^ | BioClinicalBERT | 0.836 | 0.708 | 0.613 |
|  |  | Bhate 2023 ^15^ | GPT-3.5 | 0.551 | 0.561 | 0.583 |
| Health Care Access and Quality | **Health crisis** | Scherbakov 2025 ^1^ | Mixtral 8 ×7B model | 0.77 | 0.59 | 0.48 |
|  |  | Yu 2024 ^5^ | GatorTron | 1.00 | 1.00 | 1.00 |
|  | **Insurance status** | Lituiev 2022 ^16^ | RoBERTa | - | 0.225 | - |
| Neighborhood and Built Environment | **Housing Instability** | Scherbakov 2025 ^1^ | Mixtral 8 ×7B model | 0.755 | 0.67 | 0.595 |
|  |  | Roosan 2024a ^2^ | Transformer-based | - | 0.93 | - |
|  |  | Guevara 2024 ^4^ | Flan-T5 XL + GPT-3.5 ^(b)^ | - | 0.50 | - |
|  |  | Roy 2024 ^8^ | GPT-4 | 0.62 | 0.74 | 0.92 |
|  |  | Ralevski 2024 ^23^ | GPT-4 | 0.69 | - | 0.82 |
|  |  | Yao 2023 ^24^ | Bio_ClinicalBERT | - | 0.55 | - |
|  |  | Lituiev 2022 ^16^ | RoBERTa | - | 0.627 | - |
|  |  | Han 2022 ^21^ | BERT | 0.707 | 0.552 | 0.474 |
|  |  | Gray 2023 ^18^ | ClinicalBERT | 0.74 | 0.67 | 0.64 |
|  |  | Gong 2025 ^12^ | FLAN-T5-XL + GPT turbo-0301 | - | 0.9091 | - |
|  |  | Goel 2024 ^19^ | Nous-Hermes-2-Yi-34B | - | 0.984 | - |
|  | **Living Status** | Fu 2024 ^3^ | Flan-T5-Large | - | 0.75 | - |
|  |  | Yu 2024 ^5^ | GatorTron | 0.911 | 0.914 | 0.917 |
|  |  | Ramachandran 2023 ^9^ | mSpERT | - | 0.53 | 0.526 |
|  |  | Richie 2023 ^10^ | BioClinical-BERT | 0.90 | 0.91 | 0.91 |
|  |  | Lybarger 2023 ^11^ | mSpERT | 0.80 | 0.79 | 0.77 |
|  |  | Torii 2023 ^14^ | Bio_Discharge_Summary_BERT | 0.8906 | 0.8702 | 0.8507 |
|  |  | Bhate 2023 ^15^ | GPT-3.5 | 0.568 | 0.562 | 0.583 |
|  | **Environmental Exposure** | Wang 2023 ^22^ | BioClinicalBERT | 0.954 | 0.932 | 0.911 |
|  | **Transportation** | Yu 2024 ^5^ | GatorTron | 0.911 | 0.914 | 0.917 |
|  |  | Guevara 2024 ^4^ | Flan-T5 XL + GPT-3.5 ^(b)^ | - | 0.50 | - |
|  |  | Lituiev 2022 ^16^ | RoBERTa | - | 0.622 | - |
|  |  | Han 2022 ^21^ | BERT | 0.829 | 0.482 | 0.352 |
|  |  | Gong 2025 ^12^ | FLAN-T5-XL + GPT turbo-0301 | - | 0.8571 | - |
|  |  | Gray 2023 ^18^ | ClinicalBERT | 0.88 | 0.79 | 0.72 |
| Social and Community Context | **Social Support** | Patra 2025 ^25^ | FLAN-T5-XL + GPT 4.0 (Synthetic data) | 0.79 | 0.81 | 0.86 |
|  |  | Roosan 2024a ^2^ | Transformer-based | - | 0.85 | - |
|  |  | Guevara 2024 ^4^ | Flan-T5 XL + GPT-3.5 ^(b)^ | - | 0.43 | - |
|  |  | Yu 2024 ^5^ | GatorTron | 0.976 | 0.946 | 0.926 |
|  |  | Wang 2023 ^22^ | BioClinicalBERT | 0.95 | 0.937 | 0.925 |
|  |  | Han 2022 ^21^ | BERT | 0.643 | 0.550 | 0.493 |
|  |  | Consoli 2024 ^13^ | GPT-3.5 | - | 0.9357 | - |
|  |  | Gong 2025 ^12^ | FLAN-T5-XL + GPT turbo-0301 | - | 0.6667 | - |
|  |  | Goel 2024 ^19^ | Llama-2-13b-chat-hf | - | 0.963 | - |
|  | **Bereavement** | Scherbakov 2025 ^1^ | Mixtral 8 ×7B model | 0.795 | 0.87 | 0.97 |
|  | **Interpersonal safety** | Scherbakov 2025 ^1^ | Mixtral 8 ×7B model | 0.33 | 0.50 | 1.00 |
|  |  | Yu 2024 ^5^ | GatorTron | 0.976 | 0.946 | 0.926 |
|  |  | Roy 2024 ^8^ | GPT-4 | 0.73 | 0.84 | 1.00 |
|  |  | Gabriel 2024 ^17^ | RoBERTa + GPT 3.5 turbo ^(b)^ | 1.00 | 1.00 | 1.00 |
|  |  | Botelle 2022 ^26^ | BioBERT | - | 0.95 | - |
|  | **Legal Status** | Huang 2024 ^27^ | GPT-4 | 1.00 | 0.862 | 0.757 |
|  |  | Scherbakov 2025 ^1^ | Mixtral 8 ×7B model | 0.85 | 0.89 | 0.94 |
|  |  | Roy 2024 ^8^ | GPT-4 | 0.69 | 0.73 | 0.78 |
|  |  | Goel 2024 ^19^ | Nous-Hermes-2-Yi-34B | - | 0.918 | - |
|  | **Social Isolation** | Patra 2025 ^25^ | FLAN-T5-XL + GPT 4.0 ^(b)^ | 0.82 | 0.82 | 0.82 |
|  |  | Lituiev 2022 ^16^ | RoBERTa | - | 0.636 | - |
|  | **Relationship Status** | Yu 2024 ^5^ | GatorTron | 0.976 | 0.946 | 0.926 |
|  |  | Lituiev 2022 ^16^ | RoBERTa | - | 0.772 | - |
|  |  | Gong 2025 ^12^ | FLAN-T5-XL + GPT turbo-0301 | - | 0.941 | - |
|  |  | Goel 2024 ^19^ | Nous-Hermes-2-Yi-34B | - | 0.958 | - |
| Education Access and  Quality | **Education Status** | Roosan 2024a ^2^ | Transformer-based | - | 0.87 | - |
|  |  | Fu 2024 ^3^ | Flan-T5-Large | - | 0.84 | - |
|  |  | Yu 2024 ^5^ | GatorTron | 0.967 | 0.963 | 0.959 |
|  |  | Wang 2023 ^22^ | BioClinicalBERT | 0.836 | 0.708 | 0.613 |
|  |  | Bhate 2023 ^15^ | GPT-3.5 | 0.551 | 0.561 | 0.583 |
| Behavioral | **Substance Use** | Kim 2025 ^29^ | XLM-RoBERTa + multilingual BERT | - | 0.839 | - |
|  |  | Scherbakov 2025 ^1^ | Mixtral 8 ×7B model | 0.91 | 0.95 | 1.0 |
|  |  | Rabbani 2024 ^30^ | GPT-3.5 | 0.97 | - | 0.34 |
|  |  | Shah-Mohammadi 2024a ^31^ | GPT-4 | 0.74 | 0.84 | 0.98 |
|  |  | Shah-Mohammadi 2024b ^32^ | GPT-3.5-turbo | 0.91 | 0.95 | 0.99 |
|  |  | Fu 2024 ^3^ | Flan-T5-Large | - | 0.81 | - |
|  |  | Yu 2024 ^5^ | GatorTron | 0.891 | 0.895 | 0.905 |
|  |  | Peng 2024 ^33^ | GatorTronGPT (GPT-3) | 0.842 | 0.862 | 0.862 |
|  |  | Kwon 2024 ^34^ | BioClinicalBERT | - | 0.724 | - |
|  |  | Roosan 2024b ^35^ | BERT-based | 0.89 | 0.90 | 0.92 |
|  |  | Petit-Jean 2024 ^36^ | EDS-CamemBERT | 0.972 | 0.972 | 0.972 |
|  |  | Roy 2024 ^8^ | GPT-4 | 0.868 | 0.845 | 0.83 |
|  |  | Ramachandran 2023 ^9^ | mSpERT | - | 0.611 | - |
|  |  | Turchin 2023 ^37^ | ClinicalBERT | 0.636 | 0.734 | 0.901 |
|  |  | Wang 2023 ^22^ | BioClinicalBERT | 0.956 | 0.928 | 0.902 |
|  |  | Richie 2023 ^10^ | BioClinical-BERT | 0.96 | 0.967 | 0.977 |
|  |  | Bhate 2023 ^15^ | GPT-3.5 | 0.608 | 0.636 | 0.651 |
|  |  | Kim 2023 ^38^ | XLM-RoBERTa | 0.953 | 0.847 | 0.782 |
|  |  | Kugic 2022 ^39^ | German BERT | 0.837 | 0.835 | 0.845 |
|  |  | Han 2022 ^21^ | BERT | 0.945 | 0.877 | 0.821 |
|  |  | Yu 2024 ^5^ | GatorTron | 0.891 | 0.895 | 0.905 |
|  |  | Peng 2024 ^33^ | GatorTronGPT (GPT-3) | 0.842 | 0.862 | 0.862 |
|  |  | Sajdeya 2023 ^40^ | BERT-based | 0.94 | 0.94 | 0.94 |
|  |  | Lybarger 2023 ^11^ | mSpERT | 0.853 | 0.863 | 0.873 |
|  |  | Torii 2023 ^14^ | Bio_Discharge_Summary_BERT | 0.826 | 0.793 | 0.763 |
|  | **Mental Health** | Fu 2024 ^3^ | Flan-T5-Large | - | 0.38 | - |
| (a) Best performing LLM  (b) LLM used for synthetic data generation | | | | | | |
