## Appendix C for "Beyond Metrics to Methods: A Scoping Review of Large Language Models for Detection of Social Drivers of Health in Clinical Notes"

**Framework for Methodological Assessment for LLMs Identifying SDoH in Clinical Notes**

**Introduction**

This framework provides a proposed approach to assessing the methodological rigor in developing Large Language Models (LLMs) used to identify Social Drivers of Health (SDoH) from clinical notes.

**Domain Assessment Table**

| Dimension | Domain | Assessment Question | Logical Basis for Selection |
| --- | --- | --- | --- |
| Internal Validity | **Error Analysis**^1,2^ | Were error patterns analyzed, discussed, and addressed with mitigation strategies? | Missing SDoH factors could directly harm vulnerable patients. Understanding why and how the model makes mistakes is essential to mitigate potential harm, especially when models confuse clinical data with social determinants or fail to distinguish temporary challenges from chronic social factors. Errors could lead to inappropriate resource allocation or interventions. |
|  | **Fairness Assessment**^3^ | Was performance compared across demographic groups with fairness metrics and bias mitigation? | SDoH identification must be equitable across all patient populations. Clinicians document SDoH differently across demographic groups. LLMs must recognize SDoH across different communication styles and dialects while avoiding amplifying existing biases. Performance must be consistent across race, ethnicity, gender, and socioeconomic status. |
|  | **Annotation Guidelines**^4^ | Are clear SDoH annotation guidelines provided with process documentation and quality control? | SDoH categories require clear, consistent definitions for reliable annotation. Precise definitions of what constitutes housing insecurity, food insecurity, etc. are needed. Guidelines should address how to handle indirect references to social factors, consistently rate SDoH impact severity, and distinguish current vs. historical SDoH factors. |
| External Validity | **External Validation**^5^ | Was the model validated across different datasets, demographics, time periods, and settings? | Documentation of SDoH varies dramatically between healthcare settings. Models must work equally well across demographic groups. Performance may vary based on how notes are generated (templates, free text, etc.). Different clinicians and institutions may document the same SDoH using different terminology. |
|  | **Medical Condition Specificity** | Is the medical context, healthcare setting, and relationship to SDoH clearly specified? | SDoH relevance and documentation varies by medical condition and specialty. Different medical conditions prompt different SDoH documentation. Mental health notes document different SDoH than surgical notes. Certain medical conditions have stronger connections to specific SDoH. Inpatient, outpatient, and emergency documentation capture different SDoH aspects. |
| Reporting Transparency | **Code/Prompt Availability**^6^ | Are code/prompts and model parameters available with sufficient detail for reproduction? | SDoH identification can vary dramatically based on prompt construction. Methods for adapting general LLMs to clinical text must be clear. Temperature, top-k/p settings impact SDoH identification confidence. The amount of clinical text context provided for SDoH identification is crucial. |
|  | **Dataset Availability**^6^ | Is the dataset used publicly accessible? | Dataset availability is paramount for LLMs analyzing sensitive clinical data and SDoH patterns. Clinical datasets require both privacy protections and research transparency. SDoH identification requires diverse patient populations to avoid systematic blind spots. Partial data availability may miss critical social context needed for accurate identification. Available data enables monitoring for changes in documentation practices over time. |
